# Leveraging fine-mapping and non-European training data to improve cross-population polygenic risk scores

**DOI:** 10.1101/2021.01.19.21249483

**Authors:** Omer Weissbrod, Masahiro Kanai, Huwenbo Shi, Steven Gazal, Wouter J. Peyrot, Amit V. Khera, Yukinori Okada, The Biobank Japan Project, Alicia R. Martin, Hilary Finucane, Alkes L. Price

## Abstract

Polygenic risk scores (PRS) based on European training data suffer reduced accuracy in non-European target populations, exacerbating health disparities. This loss of accuracy predominantly stems from LD differences, MAF differences (including population-specific SNPs), and/or causal effect size differences. PRS based on training data from the non-European target population do not suffer from these limitations, but are currently limited by much smaller training sample sizes. Here, we propose PolyPred, a method that improves cross-population polygenic prediction by combining two complementary predictors: a new predictor that leverages functionally informed fine-mapping to estimate causal effects (instead of tagging effects), addressing LD differences; and BOLT-LMM, a published predictor. In the special case where a large training sample is available in the non-European target population (or a closely related population), we propose PolyPred+, which further incorporates the non-European training data, addressing MAF differences and causal effect size differences. PolyPred and PolyPred+ require individual-level training data (for their BOLT-LMM component), but we also propose analogous methods that replace the BOLT-LMM component with summary statistic-based components if only summary statistics are available. We applied PolyPred to 49 diseases and complex traits in 4 UK Biobank populations using UK Biobank British training data (average *N*=325K), and observed statistically significant average relative improvements in prediction accuracy vs. BOLT-LMM ranging from +7% in South Asians to +32% in Africans (and vs. LD-pruning + P-value thresholding (P+T) ranging from +77% to +164%), consistent with simulations. We applied PolyPred+ to 23 diseases and complex traits in UK Biobank East Asians using both UK Biobank British (average *N*=325K) and Biobank Japan (average *N*=124K) training data, and observed statistically significant average relative improvements in prediction accuracy of +24% vs. BOLT-LMM and +12% vs. PolyPred. The summary statistic-based analogues of PolyPred and PolyPred+ attained similar improvements. In conclusion, PolyPred and PolyPred+ improve cross-population polygenic prediction accuracy, ameliorating health disparities.

## Introduction

Polygenic risk scores (PRS) can identify individuals at elevated risk of complex diseases, providing opportunities for preventative action^1–6^. However, many studies have shown that PRS based on European training data attain lower accuracy when applied to populations of non-European ancestry^7–26^. This loss of accuracy is primarily driven by LD differences^12–15^, allele frequency differences (including population-specific SNPs)^13,14,27^, and causal effect size differences^12–14,28–31^, though differences in heritability also play a minor role^13,14,32^. PRS based on non-European training data do not suffer from these limitations, but are currently limited by much smaller training sample sizes^1,12–15,21,33^ (however, lower non-European target sample sizes do not impact prediction accuracy). The development of new methods to reduce this gap in cross-population PRS accuracy has the potential to ameliorate health disparities^13^.

Here, we propose PolyPred, which linearly combines two complementary predictors derived from European training data: (1) PolyFun-pred, a new predictor that circumvents LD differences by applying genome-wide functionally informed fine-mapping^34,35^ to precisely estimate causal effects (instead of tagging effects); and (2) BOLT-LMM^36,37^, a published predictors that analyzes all loci jointly and can capture all signals in extremely polygenic loci. BOLT-LMM requires individual-level training data. If individual-level training data is not available, we propose two analogous methods: (i) PolyPred-S, which linearly combines PolyFun-pred with SBayesR^38^, and (ii) PolyPred-P, which linearly combines PolyFun-pred with PRS-CS^39^. Recommendations for when to use PolyPred, PolyPred-S, or PolyPred-P are provided below.

In the special case where there exists a large (e.g. *N*≥50K) non-European training sample from the target population (or a closely related population), we propose PolyPred+, a polygenic prediction method that leverages both European and non-European training data. PolyPred+ linearly combines (1) PolyFun-pred; (2) BOLT-LMM; and (3) BOLT-LMM-pop, which is obtained by applying BOLT-LMM to the non-European training data, addressing MAF differences and causal effect size differences. If individual-level training data is not available, we propose the alternative methods PolyPred-S+ and PolyPred-P+, which replace BOLT-LMM with either SBayesR or PRS-CS, respectively. Recommendations for when to use PolyPred+, PolyPred-S+, or PolyPred-P+ are provided below.

We compared PolyPred and PolyPred+ (and their summary statistic-based analogues) to state-of-the-art polygenic prediction methods via simulations and analyses of 49 diseases and complex traits in 4 populations from the UK Biobank^40^, additionally incorporating Biobank Japan^41^ and Uganda-APCDR^42,43^ to increase non-European training sample size and avoid cohort effects. We conclude that PolyPred and its summary statistic-based analogues substantially increase cross-population polygenic prediction accuracy, and that PolyPred+ and its summary statistic-based analogues further increases cross-population prediction accuracy in the special case where non-European training data is available in large sample size.

## Results

### Overview of Methods

PolyPred combines two complementary predictors: PolyFun-pred and BOLT-LMM (Table 1 and Figure 1a). PolyFun-pred is a new predictor that leverages genome-wide functionally informed fine-mapping^34,35^ to estimate posterior mean causal effects (instead of tagging effects; see Supplementary Note) for all SNPs with European MAF≥0.1% (accounting for MAF-dependent architectures^44–46^; 18 million SNPs in this study) by applying PolyFun + SuSiE^35^ to European training data across 2,763 overlapping 3Mb loci. Leveraging fine-mapped posterior mean causal effects for cross-population polygenic prediction aims to address LD differences between populations; to our knowledge, the application of PolyFun + SuSiE (or any other fine-mapping method) to polygenic prediction has not previously been explored. BOLT-LMM^36,37^ is a published predictor that estimates posterior mean tagging effects of common SNPs (1.2 million HapMap 3 SNPs^47^ in this study) using European individual-level training data. Combining PolyFun-pred with BOLT-LMM is advantageous because they have complementary advantages: PolyFun-pred estimates causal effects rather than tagging effects. BOLT-LMM estimates tagging effects, but it analyze all loci jointly, and it can potentially capture all signals in extremely polygenic loci (i.e., loci harboring >10 causal variants within 1.5Mb from the locus center; see Methods).

**Table 1:** Summary of main methods evaluated. For each method we report its constituent methods (or “-” for individual methods), the set of SNPs analyzed in model training using UK Biobank training data (and its size when restricted to imputed UK Biobank SNPs with European MAF≥0.1% and INFO score≥0.6), the training data analyzed, whether it incorporates fine-mapped effect sizes (as opposed to tagging effect sizes), whether it can work with summary statistics, and the corresponding reference. Eur: European; target pop: non-European target population; Method-pop: Method applied to training data from non-European target population.

| Method | Constituent methods | SNP set | Training data | Fine-mapped effect sizes | Summary statistics | Ref. |
| --- | --- | --- | --- | --- | --- | --- |
| P+T | - | All (18 million) | Eur | No | Yes | 48,49 |
| BOLT-LMM | - | HapMap 3 (1.2 million) | Eur | No | No | 36,37 |
| SBayesR | - | HapMap 3 (1.2 million) | Eur | No | Yes | 38 |
| PRS-CS | - | HapMap 3 (1.2 million) | Eur | No | Yes | 39 |
| PolyPred | PolyFun-pred, BOLT-LMM | All (18 million) | Eur | Yes | No | This work |
| PolyPred-S | PolyFun-pred, SBayesR | All (18 million) | Eur | Yes | Yes | This work |
| PolyPred-P | PolyFun-pred, PRS-CS | All (18 million) | Eur | Yes | Yes | This work |
| PolyPred+ | PolyFun-pred, BOLT-LMM, BOLT-LMM-pop | All (18 million) | Eur + target pop | Yes | No | This work |
| PolyPred-S+ | PolyFun-pred, SBayesR, SBayesR-pop | All (18 million) | Eur + target pop | Yes | Yes | This work |
| PolyPred-P+ | PolyFun-pred, PRS-CS, PRS-CS-pop | All (18 million) | Eur + target pop | Yes | Yes | This work |

**Figure 1:**
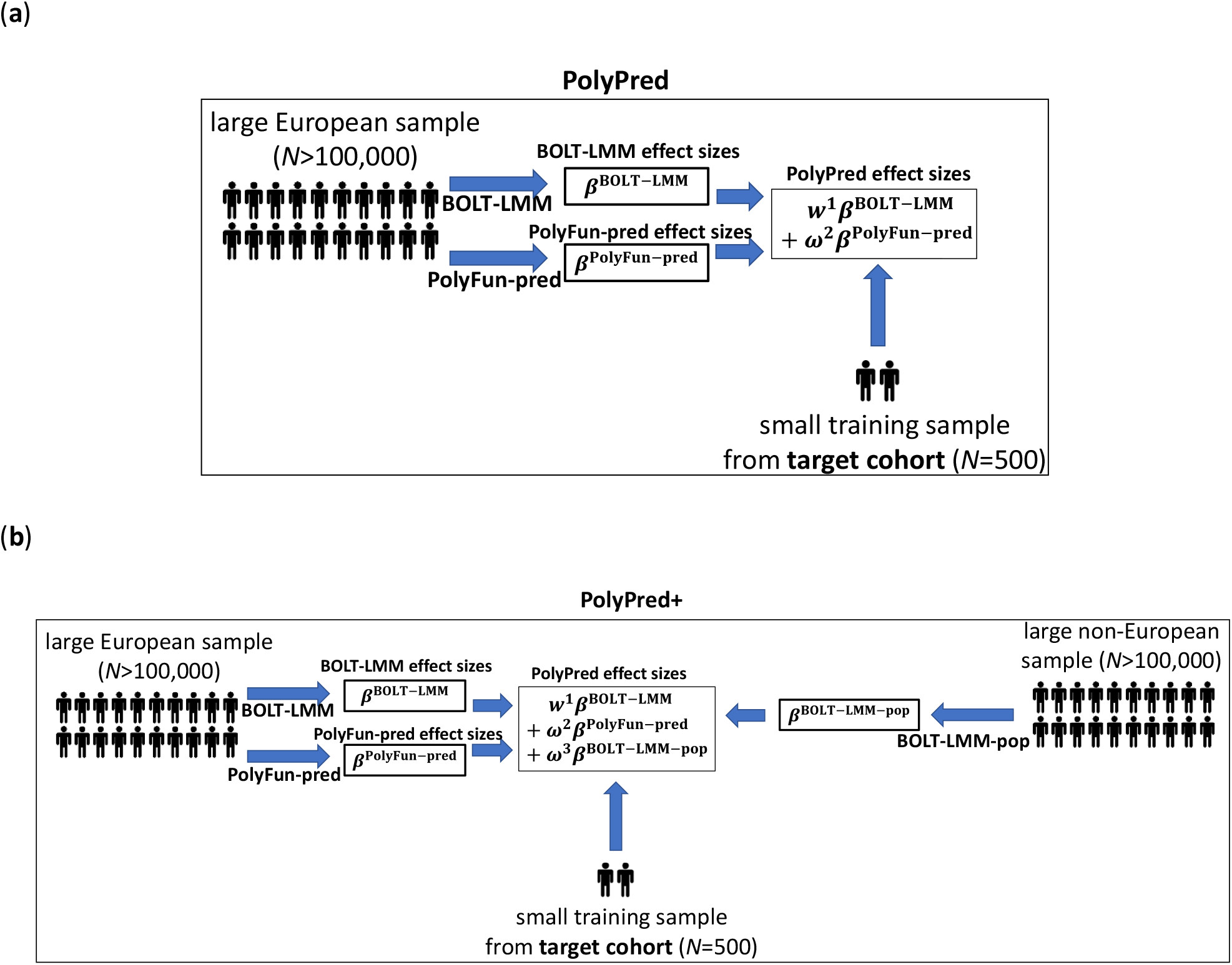
Overview of PolyPred and PolyPred+. (**a**) Overview of PolyPred. PolyPred linearly combines the effect sizes of BOLT-LMM (*β*^BOLT−LMM^) and PolyFun-pred (*β*^PolyFun−pred^), (trained using European training data). It uses a small training sample from the target population to estimate mixing weights (*ω*^1^, *ω*^2^) for the constituent methods. (**b**) Overview of PolyPred+. PolyPred+ linearly combines the effect sizes of BOLT-LMM (*β*^BOLT−LMM^), PolyFun-pred (*β*^PolyFun−pred^) (trained using European training data), and BOLT-LMM-pop (*β*^BOLT−LMM−pop^) (trained using non-European training data from the target population). It uses a small training sample from the target population to estimate mixing weights (*ω*^1^, *ω*^2^, *ω*^3^) for the constituent methods. PolyPred-S and PolyPred-P (resp. Poly-Pred-S+ and PolyPred-P+) replace all instances of BOLT-LMM with SBayesR or PRS-CS, respectively.

In the special case where a large training sample is available in the target population (or a closely related population), we propose PolyPred+, which combines three complementary predictors: PolyFun-pred, BOLT-LMM, and BOLT-LMM-pop (Table 1 and Figure 1b); BOLT-LMM-pop refers to application of BOLT-LMM to common SNPs (1.2 million HapMap 3 SNPs in this study) using training data from the non-European target population, addressing MAF differences and causal effect size differences.

PolyPred computes linear combinations of the estimated effect sizes of their constituent predictors:

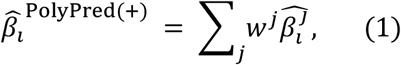

where *i* indexes SNPs, *j* indexes the constituent predictors (PolyFun-pred and BOLT-LMM for PolyPred; PolyFun-pred, BOLT-LMM and BOLT-LMM-pop for PolyPred+), 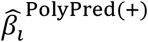 is the PolyPred (+) per-allele effect size of SNP *i, w*^*j*^ are method-specific weights, and 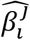 is the per-allele effect size of SNP *i* for method *j* (or 0 if SNP *i* was not considered by method *j*). Predicted phenotypes are computed by applying effect sizes to target genotypes:

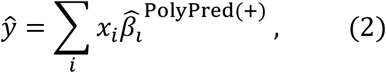

where *ŷ* is the predicted phenotype of an individual from the target population and *x*_*i*_ is the number of minor alleles of SNP *i* carried by the individual. The mixing weights *w*^*j*^ in Equation 1 are estimated via non-negative least squares regression using a small number of training individuals from the target population (500 in this study), regressing true phenotypes on a linear combination of the constituent predictors (which are computed as in Equation 2).

PolyPred requires individual-level training data for its BOLT-LMM component. If only summary statistics (and summary LD information) are available, we propose two analogous methods (Table 1): (i) PolyPred-S, which linearly combines PolyFun-pred and SBayesR^38^; and (ii) PolyPred-P, which linearly combines PolyFun-pred and PRS-CS^39^. We also propose the analogous methods PolyPred-P+ and PolyPred-S+ (Table 1). Further details of PolyPred and PolyPred+ (and their summary statistic-based analogues) are provided in the Methods section; we have publicly released open-source software implementing these methods (see URLs).

We evaluate prediction accuracy for each method and target population using relative-*R*^2^, defined as the *R*^2^ obtained in the target non-European population (after correcting for covariates and potential confounders; see Methods) divided by the *R*^2^ obtained by BOLT-LMM in UK Biobank non-British Europeans (employing the same correction), using the same training data for the numerator and the denominator. This quotient transforms the prediction accuracies from an absolute scale to a scale of relative improvement (vs. the BOLT-LMM predictor in the UK Biobank non-British European target population), which is invariant to factors such as training sample size and trait heritability. We compute standard errors via a genomic block-jackknife, which is conservative compared to a jackknife over individuals (see Methods). We meta-analyze relative-*R*^2^ across traits in each target population via an inverse variance-weighted average, weighting traits according to the sampling variance of the BOLT-LMM predictor in the target population (estimated via genomic block-jackknife; see Methods). We compare PolyPred and PolyPred+ (and their summary statistic-based analogues) to 4 published methods: LD-pruning + P-value thresholding (P+T)^48,49^, BOLT-LMM^36,37^, SBayesR^38^, and PRS-CS^39^ (Table 1).

Our recommendation for which version of PolyPred to use (see Table 1) depends on three main factors: (i) whether individual-level training data is available; (ii) the size and consistency of matched ancestry of the LD reference panel (if individual-level training data is not available); and (iii) whether non-European training data is available. Our results for the underlying constituent methods are summarized in Table 2 (detailed below), and our recommendations are summarized in Figure 2.

**Table 2:** Summary of the relative performance of constituent PRS methods. For each of three constituent PRS methods (BOLT-LMM, SBayesR, PRS-CS) we report its relative performance in prediction in UK Biobank non-British Europeans under various settings; we also provide links to the corresponding Figure(s)/Table(s) (red font for simulations, blue font for real trait analyses). **✓✓**: the method is significantly more accurate than the second best method in the same row, and combining this method with PolyFun-pred increases prediction accuracy; **✓✓**^*^: the method is significantly more accurate than the second best method in the same row, and combining this method with PolyFun-pred does not increase prediction accuracy; **✓**: the method is significantly less accurate than the best method in the same row, but is significantly more accurate than P+T; **✗**: the method is not significantly more accurate than P+T; **---**: the method is not applicable, because it requires individual-level data. For Very large unmatched LD (a likely scenario when analyzing summary statistics from a meta-analysis of many cohorts), we performed real trait analyses only, as simulations would have required another very large individual-level data set in addition to UK Biobank (see Supplementary Note). For Individual-level data, the difference between BOLT-LMM and the second-best method was significant in simulations but non-significant in real trait analyses. For In-sample LD, the difference between SBayesR and PRS-CS was significant in simulations but non-significant in real traits analyses. For Very large unmatched LD (a likely scenario when analyzing summary statistics from a meta-analysis of many cohorts), we performed real trait analyses only (see explanation in Supplementary Note). For small unmatched LD, we performed both simulations and real trait analyses but report results of real trait analyses, which we believe to be most reflective of real-life settings (in simulations, SBayesR was significantly more accurate than PRS-CS). Results for non-European target populations from UK Biobank were similar, though some of the differences were not statistically significant due to smaller prediction accuracies and sample sizes. We have facilitated the use of very large LD reference panels for European training data by publicly releasing summary LD information for *N*=337K British-ancestry UK Biobank samples across 18 million SNPs (see Data availability).

| LD | BOLT-LMM | SBayesR | PRS-CS | Figure(s)/Table(s) |
| --- | --- | --- | --- | --- |
| Individual-level data (UKB, $N=337K$) | ✓✓ | ✓ | ✓ | Figures 3,4,7 |
| In-sample LD (UKB, $N=337K$) | --- | ✓✓ | ✓ | Figures 3,4,7 |
| Very large unmatched LD (UKB, $N=337K$) | --- | ✓ | ✓✓ | Figure 5 |
| Small unmatched LD (1000G, $N=489$) | --- | ✗ | ✓✓* | Tables S4-S6 |

**Figure 2:**
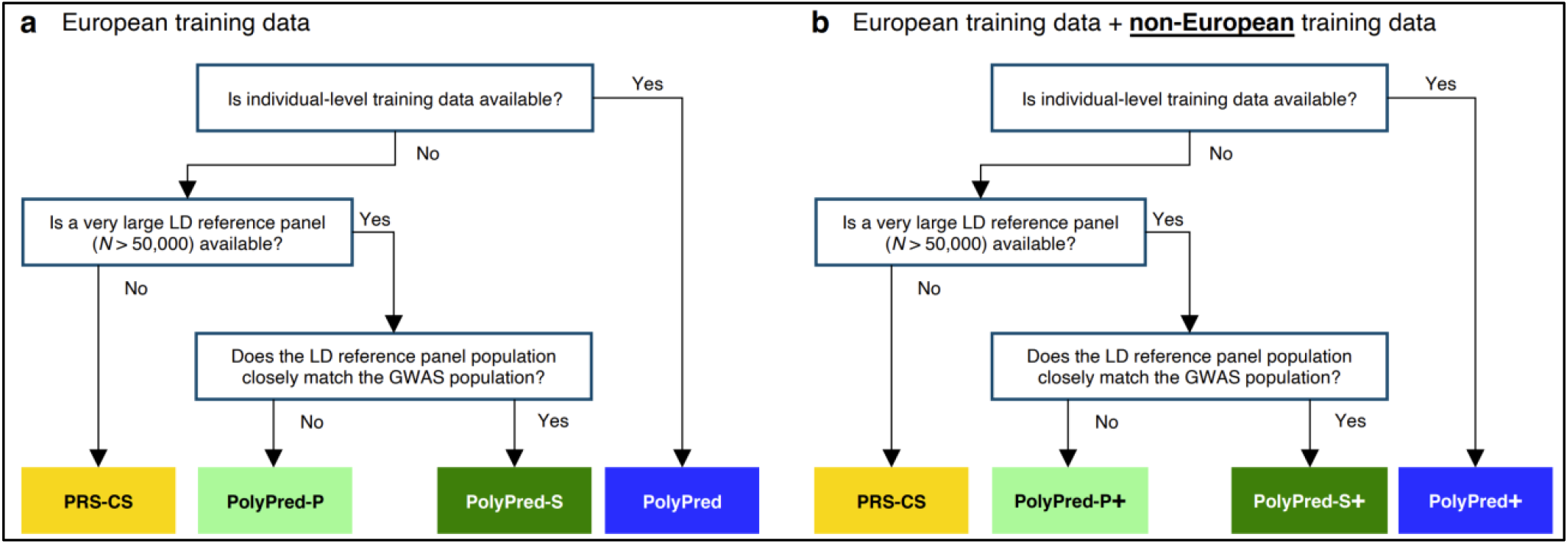
Recommendations for the application of PolyPred, PolyPred+ and related methods. (**a**) Flowchart of recommendations when only European training data is available. (**b**) Flowchart of recommendations when both European and non-European training data are available. We note that when working with summary statistics from a meta-analysis of many cohorts, there is typically no LD reference panel that closely matches the GWAS population. Also, it is possible that the answers to the flowchart questions are different for European vs. non-European training data, in which case the recommendation would be to use a hybrid method based on the answers to each flowchart in turn (e.g. PolyFun-pred + BOLT-LMM + PRS-CS-pop; not listed in Table 1). For both (a) and (b), we recommend training PolyFun-pred using a very large LD reference panel (e.g. *N*=337K UK Biobank British) with a dense SNP set (e.g. 8 million SNPs). We have facilitated this by publicly releasing summary LD information for *N*=337K British-ancestry UK Biobank samples across 18 million SNPs (see Data availability).

### Simulations with in-sample LD

We compared PolyPred, PolyPred-S and PolyPred-P to P+T, BOLT-LMM, SBayesR, and PRS-CS via simulations, using real genotypes or in-sample LD (i.e. LD data based on the GWAS sample) from the UK Biobank^40^. We trained each method using 337,491 unrelated British-ancestry individuals^40^, and computed predictions in four target populations: non-British Europeans, South Asians, East Asians, and Africans. We estimated mixing weights for PolyPred, PolyPred-S and PolyPred-P using 500 individuals from the target population. We evaluated prediction accuracy using held-out individuals from each target population that were not included in the training sets: 42K non-British Europeans, 7.7K South Asians, 0.9K East Asians, and 6.2K Africans. We computed PRS using 250,963 MAF≥0.1% SNPs with INFO score≥0.6 on chromosome 22 (including short indels) (we restricted the analysis to chromosome 22 due to alleviate the computational burden of running hundreds of simulations). Generative trait architectures were specified as follows. We simulated traits with polygenicity (genome-wide proportion of causal SNPs) equal to either 0.1% (less polygenic) or 0.3% (more polygenic) and heritability equal to 5% (we specified a heritability that is larger than typical chromosome 22 heritability to increase our power to detect differences between methods using a limited number of simulations; see below). We specified prior causal probabilities for each SNP in proportion to per-SNP heritabilities, which we generated for each SNP based on its British LD, MAF, and functional annotations, using the baseline-LF model^45^ with meta-analyzed functional enrichments from real traits as described in our previous work^35^, and sampled causal SNPs. For each causal SNP, we sampled ancestry-specific causal effect sizes (for European, South Asian, East Asian, and African ancestries) from a multivariate normal distribution assuming cross-population genetic correlations of 0.8, consistent with recent findings^13,30^; functional annotations impacted prior causal probabilities but not causal effect sizes for causal SNPs, consistent with our recent work^50^. Other parameter settings were explored in secondary analyses (see below). Further details of the simulation framework are provided in the Methods section.

We computed summary association statistics (used by every method except BOLT-LMM) via linear regression. For SBayesR, PRS-CS, and PolyFun-pred, we computed summary LD from a very large (*N*≥50K) subset of the UK Biobank British population, effectively using in-sample LD: For SBayesR we used summary LD for 18,040 HapMap 3 SNPs on chromosome 22 estimated from 50K British-ancestry UK Biobank individuals, that was made publicly available by the authors of SBayesR^51^; for PRS-CS we used summary LD for 16,214 HapMap 3 SNPs on chromosome 22 estimated from 375,120 British-ancestry UK Biobank individuals, which was made publicly available (in the form required by PRS-CS) by the authors of PRS-CS^39^; and for PolyFun-pred we used summary LD estimated from 337,548 British-ancestry UK Biobank individuals that we previously made publicly available^35^. For P+T, we used summary LD estimated from a random subset of 10,000 British-ancestry UK Biobank individuals to alleviate computational costs. For BOLT-LMM, we used individual-level genotypes at 18,040 HapMap 3 SNPs on chromosome 22, using hard-called values for imputed alleles. We applied all methods using default or recommended parameter settings (Methods). We computed relative-*R*^2^ for each method, target population, and trait architecture (less polygenic, more polygenic), averaged across 100 simulations. We did not evaluate PolyPred+, PolyPred-S+ and PolyPred-P+ in these experiments because of the small size of the UK Biobank non-European populations. In addition to the simulations with in-sample LD described below, we also performed simulations with reference panel LD (Supplementary Note; also see Table 2).

The simulation results are reported in Figure 3 and Supplementary Table 1 (also see Table 2). PolyPred was the most accurate method in each target population for both trait architectures, with relative improvements vs. BOLT-LMM (resp. P-values for improvement) ranging from +13% in non-British Europeans (P<10^−16^) to +65% in Africans (P<10^−16^) for the less polygenic architecture, and from +2% in non-British Europeans (P=0.0001) to +17% in Africans (P=10^−8^) for the more polygenic architecture. PolyPred-S and PolyPred-P performed slightly worse than PolyPred for both trait architectures, but were substantially and significantly more accurate than the corresponding constituent methods (SBayesR for PolyPred-S, PRS-CS for PolyPred-P). Among the remaining methods, BOLT-LMM was consistently the most accurate and P+T was consistently the least accurate method, far underperforming the other methods (despite its widespread recent use^11,13–18,23,31,52–56^). We note that the higher accuracy of BOLT-LMM vs. SBayesR and PRS-CS does not imply that BOLT-LMM is a superior method, as BOLT-LMM analyzes individual-level training data whereas SBayesR and PRS-CS analyze summary statistics (there also exist other methods that analyze individual-level training data, e.g. BayesR^57^). We emphasize that although concentrating 5% heritability into chromosome 22 increases absolute *R*^2^, this is not expected to impact relative-*R*^2^ (or relative improvements vs. BOLT-LMM).

**Figure 3:**
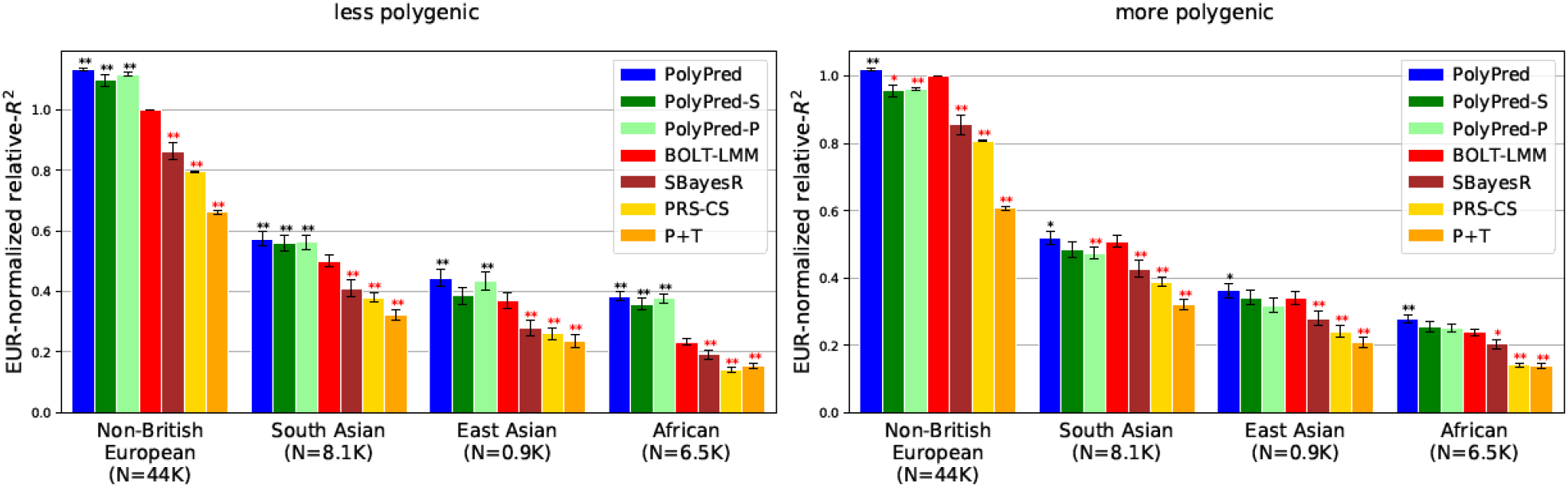
Cross-population PRS results for simulated UK Biobank traits using in-sample LD. We report average prediction accuracy (relative-*R*^2^; see main text) for PRS trained in UK Biobank British samples (*N*=337K) and applied to 4 UK Biobank target populations across 100 simulated traits with less polygenic (0.1% of SNPs causal; left panel) or more polygenic (0.3% of SNPs causal; right panel) architectures. Target population sample sizes are indicated in parentheses; PolyPred and its summary statistic-based analogues used 500 additional training samples from each target population to estimate mixing weights. Asterisks above each bar denote statistical significance of the difference vs. BOLT-LMM, with black asterisks denoting an advantage and red asterisks denoting a disadvantage (*P<0.05; **P<0.001). Errors bars denote standard errors. Numerical results, absolute prediction accuracies (*R*^2^), and P-values of relative improvements vs. BOLT-LMM are reported in Supplementary Table 1.

We performed 5 secondary analyses to investigate the sensitivity of the results to the simulation parameters. First, we performed simulations for much less polygenic (0.05%) and much more polygenic (0.5%) architectures. PolyPred remained the most accurate method, attaining the largest relative improvements vs. BOLT-LMM for the much less polygenic architecture, with slightly worse results for PolyPred-S and PolyPred-P (Supplementary Table 1); we conservatively restricted the remaining secondary analyses to the more polygenic (0.3%) architecture (for which PolyPred attains smaller relative improvements among the two main architectures simulated) and omitted PolyPred-S and PolyPred-P (due to their close similarity to PolyPred), unless otherwise indicated. Second, we performed simulations with lower (3%) or higher (7%) chromosome 22 heritability. PolyPred remained the most accurate method, with relative improvements vs. BOLT-LMM increasing with heritability (Supplementary Table 1). Third, we performed simulations with cross-population genetic correlations increased from 0.8 to 1.0. PolyPred remained the most accurate method, with relative improvements vs. BOLT-LMM remaining broadly similar (Supplementary Table 1). Fourth, we modified the number of training samples from the target population used to estimate mixing weights (*N*_mix_) from 500 to various values from 100-1000. PolyPred remained the most accurate method in all these experiments, with relative improvements vs. BOLT-LMM increasing with *N*_mix_ but limited improvement above *N*_mix_=500 (Supplementary Table 1). Fifth, we decreased the number of British-ancestry training samples (*N*) from *N*=337K to *N*=100K or *N*=10K. Prediction accuracies decreased with decreasing training sample size for all methods, and the relative improvements of PolyPred vs. BOLT-LMM (and other methods) were substantially decreased for *N*=10K, though they remained statistically significant in Africans under 0.1% polygenicity (Supplementary Table 1).

We performed two secondary analyses to investigate the sensitivity of the results to the SNP set and functional annotations. First, we evaluated a modified version of PolyPred that uses only 1.2 million HapMap 3 SNPs (matching the SNP sets of BOLT-LMM, SBayesR, and PRS-CS) instead of 18 million SNPs. PolyPred suffered a substantial loss of accuracy in this setting, demonstrating the importance of using a dense SNP set for fine-mapping based PRS (Supplementary Table 1). Second, we evaluated a non-functionally informed method (PolyPred-NoFun) that linearly combines PolyNoFun-pred (a modification of PolyFun-pred that is not functionally-informed; see Methods) and BOLT-LMM, precluding the need for functional annotations. PolyPred-NoFun was slightly less accurate than PolyPred, but still more accurate than BOLT-LMM (Supplementary Table 1).

We performed two secondary analyses to evaluate the computational cost and memory cost of each method. First, we evaluated the computational cost of each method (for PolyPred, PolyPred-S, and PolyPred-P, we included the time cost of each constituent method); we focused on the time cost to compute SNP effect sizes used for prediction, as the time cost to compute predictions in target samples using these SNP effect sizes is approximately the same for each method. SBayesR was the fastest method (2.8 minutes), P+T was the second fastest method (7.4 minutes), PRS-CS was the third fastest method (113 minutes), BOLT-LMM was the fourth fastest method (224 minutes), PolyPred-S was the fifth fastest method (447 minutes), PolyPred-P was sixth fastest method (557 minutes), and PolyPred was the slowest method (668 minutes) (Supplementary Table 2). Second, we evaluated the memory cost of each method (for PolyPred, we computed the maximum memory cost of each constituent method). We performed this analysis using chromosome 1 instead of chromosome 22 because memory cost can increase with the number of SNPs in the analysis (but the memory cost of PolyFun-pred is fixed because it analyzes each 3Mb-locus separately). P+T used the least memory (1.5GB), PRS-CS used the second smallest amount of memory (1.8GB), SBayesR used the third smallest amount of memory (2.6GB), BOLT-LMM used the fourth smallest amount of memory (11GB), and PolyPred, PolyPred-S, and PolyPred-P all used the most memory (57GB) (Supplementary Table 2). The larger computational cost of PolyPred and its summary statistic-based analogues is dominated by the PolyFun-pred component, which is computationally intensive because (i) it performs fine-mapping and (ii) it analyses a large number of SNPs (see Discussion).

We conclude that PolyPred and its summary statistic-based analogues are more accurate than BOLT-LMM, SBayesR, PRS-CS, and P+T, with small but significant improvements vs. BOLT-LMM in Europeans and substantial improvements in Africans.

### Analysis of 4 UK Biobank populations using UK Biobank British training data

We applied PolyPred and its summary statistic-based analogues to 49 diseases and complex traits from the UK Biobank, analyzing 4 target populations (Methods, Supplementary Table 3). As in our simulations, we used UK Biobank British training data (average *N*=325K) to estimate SNP effect sizes; used 500 additional individuals from the target population to estimate mixing weights (we note that PolyPred and its summary statistic-based analogues are relatively insensitive to the choice of mixing weights; see below); evaluated prediction accuracy using individuals from each of the 4 target populations that were not included in the training data, and were unrelated to the training individuals and to each other: 42K non-British Europeans, 7.7K South Asians, 0.9K East Asians, and 6.2K Africans; and compared PolyPred and its summary statistic-based analogues to P+T, BOLT-LMM, SBayesR, and PRS-CS. We meta-analyzed relative-*R*^2^ across traits by restricting to 7 well-powered, independent complex traits from the UK Biobank^40^ (|*r*_g_|<0.3; see Methods and Supplementary Table 3) that were also available in Biobank Japan and in Uganda-APCDR (see below). We excluded the HLA region and two other long-range LD regions from the analysis (Methods). We have publicly released SNP effect sizes used for prediction for each of the 4 methods (see URLs).

We computed relative-*R*^2^ for each method and target population. Results meta-analyzed across traits are reported in Figure 4 and Supplementary Tables 4-6 (also see Table 2), and results for each trait are reported in Supplementary Tables 4-6. Among the published methods, BOLT-LMM attained the highest prediction accuracy in all target populations (differences between BOLT-LMM and SBayesR were small and not statistically significant, but the difference between BOLT-LMM and PRS-CS was statistically significant in non-British Europeans); as noted above, the higher accuracy of BOLT-LMM vs. SBayesR and PRS-CS does not imply that BOLT-LMM is a superior method, as BOLT-LMM analyzes individual-level training data whereas SBayesR and PRS-CS analyze summary statistics. P+T was much less accurate than the other methods (despite its widespread recent use^11,13–18,23,31,52–56^), suffering relative losses of 37-50% vs. BOLT-LMM. We thus used BOLT-LMM as a benchmark, conservatively assessing the statistical significance of improvements vs. BOLT-LMM via genomic block-jackknife across 200 genomic regions (Methods).

**Figure 4:**
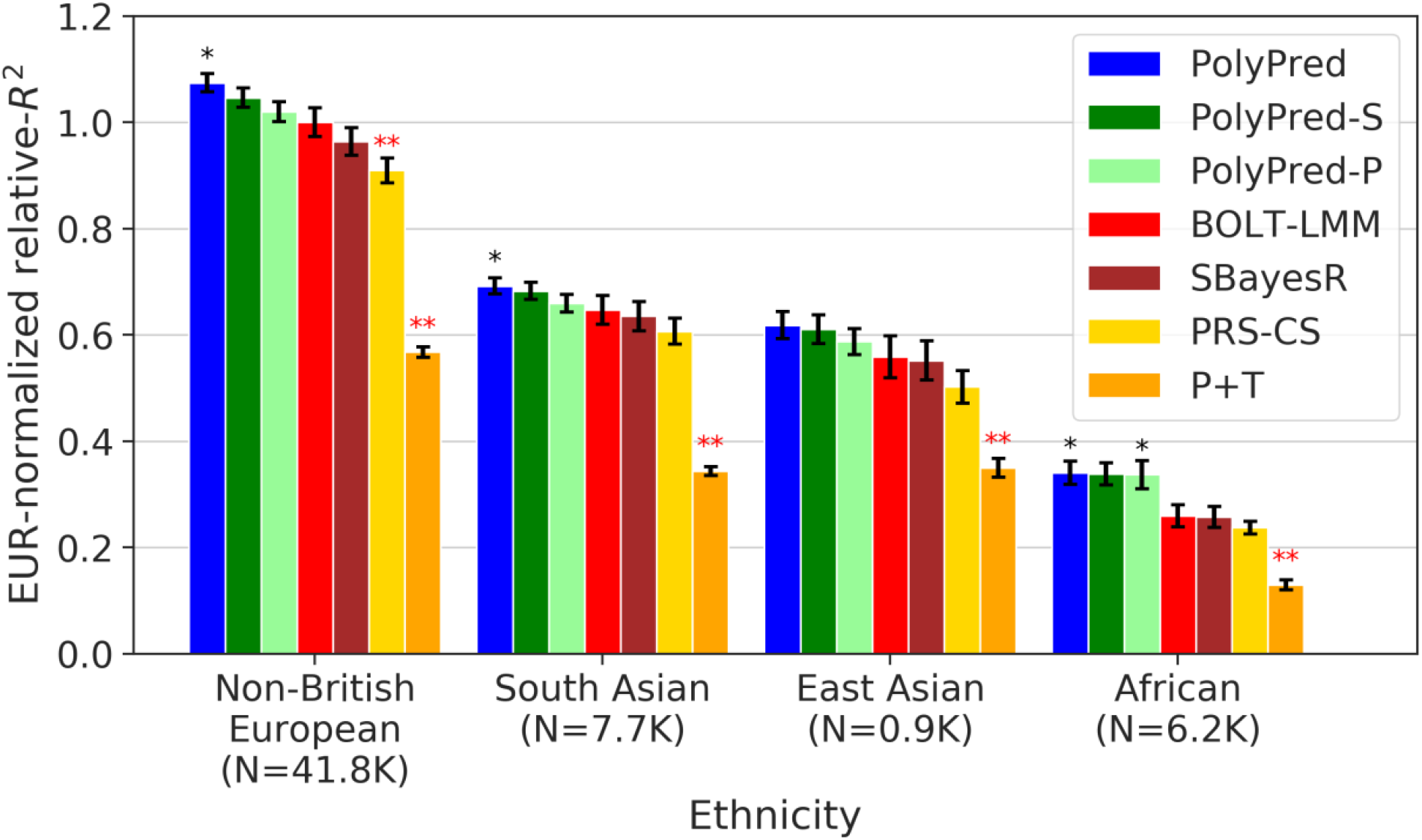
Cross-population PRS results for real UK Biobank traits. We report average prediction accuracy (relative-*R*^2^; see main text), meta-analyzed across 7 well-powered, independent traits, for PRS trained in UK Biobank British samples (average *N*=325K) and applied to 4 UK Biobank target populations. Target population sample sizes are indicated in parentheses; PolyPred and its summary statistic-based analogues used 500 additional training samples from each target population to estimate mixing weights. Asterisks above each bar denote statistical significance of the difference vs. BOLT-LMM, with black asterisks denoting an advantage and red asterisks denoting a disadvantage (*P<0.05; **P<0.001). Errors bars denote standard errors. Numerical results, results for all 49 traits analyzed, absolute prediction accuracies (*R*^2^), and P-values of relative improvements vs. BOLT-LMM are reported in Supplementary Tables 4-6.

Among all 7 methods, PolyPred attained the highest prediction accuracy in each target population. Improvements in average relative-*R*^2^ of PolyPred vs. BOLT-LMM were equal to +7.5% in non-British Europeans (P=0.05), +6.8% in South Asians (P=0.02), +11% in East Asians (P=0.12) and +32% in Africans (P=0.02). The larger improvement in Africans reflects the larger LD differences vs. British training data, due to earlier divergence times^13,14,58^. The lack of statistical significance in East Asians reflects the low power to detect significant differences in very small target samples (though statistical power is primarily limited by genome size due to our conservative use of genomic block-jackknife). PolyPred-S was consistently the second most accurate method, with statistically significant improvements vs. SBayesR (and no statistically significant differences vs. PolyPred; up to 3% reduction in average relative-*R*^2^). PolyPred-P was consistently the third most accurate method, with a statistically significant improvement vs. PRS-CS (and a statistically significant differences vs. PolyPred in Non-British Europeans; up to 5% reduction in average relative-*R*^2^). The relative mixing weight contributions of PolyFun-pred/BOLT-LMM to PolyPred were equal to 38%/72% in non-British Europeans, 40%/60% in South Asians, 63%/37% in East Asians, and 48%/52% in Africans (Methods, Supplementary Table 4). Despite the improvements attained by PolyPred, the reductions in prediction accuracy in non-European populations remained substantial, with meta-analyzed absolute *R*^2^ equal to 0.17 in non-British Europeans, 0.11 in South Asians, 0.093 in East Asians, and 0.053 in Africans. These reductions were highly statistically significant (P<0.002 for all analyses involving European-ancestry training data) (Methods, Supplementary Tables 4-5).

We assessed the calibration of each prediction method. A predictor is correctly calibrated if a regression of the true phenotype vs. the predictor yields a slope of 1, and is miscalibrated otherwise^33^. Regression slopes are reported in Supplementary Table 4. In non-British Europeans, PolyPred was well-calibrated (regression slope = 1.01), BOLT-LMM and SBayesR were approximately well-calibrated (0.96-1.08), PRS-CS was slightly miscalibrated (1.26), and P+T was poorly calibrated (0.08). In non-European populations, PRS-CS was approximately well-calibrated (0.85-1.11), but BOLT-LMM and SBayesR suffered reduced regression slopes (0.57-0.90), consistent with reduced prediction accuracy. In contrast, PolyPred and its summary statistic-based analogues remained well-calibrated (0.95-1.17), as expected due to their extra training step to estimate mixing weights in the target population. For brevity, we focus on PolyPred (instead of its summary statistic-based analogues) in the remainder of this subsection (results for all methods are reported in Supplementary Tables 4-6).

We performed 5 secondary analyses to evaluate the impact of the LD reference panel and the SNP set on prediction accuracy (we note that analyses of summary statistics from a meta-analysis of many cohorts generally require using an LD reference panel instead of in-sample LD). First, we evaluated a modified version of PolyFun-pred using a reference panel based on UK10K (*N*=3,567), and observed a substantial and statistically significant reduction in accuracy, to a far greater degree that observed in simulations (Supplementary Tables 4-6). Second, we evaluated a modified version of PRS-CS that uses an LD reference panel from 1000 Genomes project Europeans (*N*=489) and observed statistically indistinguishable results from those obtained using in-sample LD (unlike in simulations, where we observed significantly reduced accuracy when using an LD reference panel from 1000 Genomes project Europeans) (Supplementary Tables 4-6). Third, we evaluated modified versions of SBayesR that use (i) an LD reference panel using UK10K (*N*=3,567); (ii) an LD reference panel using 1000 Genomes project Europeans (*N*=489); or (iii) an LD reference panel using a subset of UK10K (*N*=489). We observed (i) very similar and statistically indistinguishable accuracy when using UK10K, (ii) severely reduced accuracy (P<4×10^−6^) when using 1000 Genomes project Europeans, and (iii) moderately reduced accuracy (P=0.07 in East-Asians, P<7×10^−6^ in other target populations) when using a subset of UK10K, suggesting that the loss of accuracy primarily stems from LD mismatch rather than reduced sample size (Supplementary Tables 4-6). Fourth, we evaluated a modified version of SBayesR (SBayesR-2.8M) that uses 2.8M common SNPs specified by the authors of SBayesR^38^ instead of 1.2 million HapMap 3 SNPs. SBayesR-2.8M was less accurate than SBayesR (significantly so for Africans) (Supplementary Tables 4-6). Thus, our use of SBayesR (using 1.2 million HapMap 3 SNPs) instead of SBayesR-2.8M in all primary comparisons is a conservative choice, since SBayesR outperforms SBayesR-2.8M (we note that naively scaling SBayesR and PRS-CS to use 18 million SNPs as in PolyFun-pred would be computationally infeasible^38,39^). Fifth, we evaluated a modified version of BOLT-LMM (BOLT-LMM-727K) that estimates effect sizes using only 727K genotyped SNPs (instead of 1.2 million imputed HapMap 3 SNPs). BOLT-LMM-727K was substantially and significantly less accurate than BOLT-LMM (Supplementary Table 4).

We performed 10 additional secondary analyses. First, we evaluated LDpred^33^ using 1000 Genomes project Europeans^59^ or UK10K^60^ as the LD reference panel (Methods). Both versions of LDpred were consistently less accurate than BOLT-LMM (Supplementary Table 4). Second, we evaluated modified versions of PolyPred that specify fixed mixing weights instead of estimating mixing weights in the target populations. We considered mixing weights for PolyFun-pred/BOLT-LMM equal to 0%/100%, 25%/75%, 50%/50%, 75%/25%, and 100%/0%. The 25%/75% and 50%/50% methods performed very similarly to PolyPred, with no statistically significant differences (Supplementary Table 6). Third, we restricted the PolyFun-pred component of PolyPred to only include SNPs with posterior causal probability greater than a fixed threshold (0.05, 0.50 or 0.95). This restriction decreased prediction accuracy (Supplementary Table 4,6), implying that estimating causal effect sizes is beneficial for prediction even at loci that cannot be confidently fine-mapped. Fourth, we evaluated a non-functionally informed method (PolyPred-NoFun) that linearly combines PolyNoFun-pred (a modification of PolyFun-pred that is not functionally-informed; see Methods) and BOLT-LMM. PolyPred-NoFun was slightly less accurate than PolyPred, but still more accurate than BOLT-LMM (Supplementary Tables 4,6). The difference between PolyPred-NoFun vs. PolyPred was not statistically significant, in contrast to previous studies reporting a large and statistically significant increase in prediction accuracy from incorporating functional annotations^61–63^. Fifth, we reduced the number of training samples from the target population used to estimate mixing weights (*N*_mix_) from 500 to 100. PolyPred suffered slightly reduced accuracy but remained the most accurate method, although relative improvements vs. BOLT-LMM were no longer statistically significant due to larger standard errors (Supplementary Table 4). Sixth, we computed standard errors of relative-*R*^2^ using a jackknife over individuals^61^ (instead of a genomic block-jackknife over SNPs; see Methods). Standard errors computed using a jackknife over individuals were generally smaller, increasing the statistical significance of relative improvements of PolyPred vs. BOLT-LMM (Supplementary Table 4). Seventh, we meta-analyzed the results of each method across three independent diseases: type 2 diabetes, asthma, and all autoimmune disease (Methods); these diseases were not included in our primary meta-analyses due to low (observed-scale) heritabilities. PolyPred attained the highest prediction accuracy for each target population and each disease, except for East Asians (where standard errors were very large in relative terms due to the small sample size) and for type 2 diabetes in non-British Europeans (where BOLT-LMM performed slightly but non-significantly better) (Supplementary Table 4). However, relative improvements were not statistically significant due to lower power (Supplementary Table 4). Eighth, we observed very similar results when down-sampling the non-British European target sample size to match the African target sample size, demonstrating that the reduced accuracy in Africans vs. Europeans is not due to the lower target sample size (Supplementary Table 4). Ninth, we evaluated two versions of PRS-CS that use pre-specified values of its global shrinkage parameter (0.01 and 0.001, following the recommendations of the authors of PRS-CS^39^). Both versions were less accurate than the default version of PRS-CS (which automatically adjusts the value of this parameter), justifying the use of the default version of PRS-CS in this work (Supplementary Tables 4-5). Finally, we assessed the potential contribution of ancestry-specific heritability to reductions in cross-population prediction accuracy^14^, by applying GCTA^64^ to estimate the SNP-heritability explained by HapMap 3 SNPs^65,66^ in each target population. SNP-heritabilities were largest in non-British Europeans and smallest in Africans (Supplementary Table 7) (these differences could be due to SNP ascertainment^67^, sample ascertainment, and/or ancestry-specific architectures^30^), likely contributing to reductions in cross-population prediction accuracy.

We conclude that PolyPred and its summary statistic-based analogues substantially increase cross-population polygenic prediction accuracy vs. published methods (with a particularly large improvement in Africans), consistent with simulations. However, there remains a large gap in cross-population polygenic prediction accuracy as compared to Europeans.

### Analysis of 4 UK Biobank populations using ENGAGE meta-analysis training data

We sought to analyze training data consisting of summary statistics for real traits from a meta-analysis of many European cohorts, for which in-sample LD is generally not available. We analyzed 8.1 million meta-analyzed summary statistics from the European Network for Genetic and Genomic Epidemiology (ENGAGE) consortium^68–70^ for four traits (BMI, waist-hip-ratio (adjusted for BMI), total cholesterol, and triglycerides; average *N*=61,365), and evaluated the prediction accuracy using the same four UK Biobank populations analyzed previously (Non-British Europeans, South-Asians, East-Asians, and Africans). We selected this particular meta-analysis because it includes a dense set of 8.1 million imputed SNPs, which enables fine-mapping. For each method, we used an LD reference panel based on UK Biobank British individuals (*N*=337K for PolyFun-pred and PRS-CS, *N*=50K for SBayesR), as previously described; we emphasize that unlike the other primary analyses in this manuscript, the LD reference panel was mis-specified, because it was not based on in-sample LD. We excluded methods that require individual-level training data (BOLT-LMM and PolyPred) from this analysis.

Results meta-analyzed across traits are reported in Figure 5, Supplementary Table 5, and Supplementary Table 8 (also see Table 2), and results for individual traits are reported in Supplementary Table 5 and Supplementary Table 8. Briefly, PolyPred-P was generally the most accurate method, and PRS-CS outperformed SBayesR (with a significant improvement for non-British Europeans and Africans), consistent with a previous study^71^ (unlike our analysis of UK Biobank training data, where SBayesR outperformed PRS-CS; Figure 4). In detail, the average relative-*R*^2^ in Non-British Europeans was 0.045 for PolyPred-P, 0.044 for PolyPred-S, 0.039 for PRS-CS, 0.033 for SBayesR, and 0.022 for P+T. In Africans, the average relative-*R*^2^ was 0.015 for PolyPred-P, 0.008 for PolyPred-S, 0.013 for PRS-CS, 0.010 for P+T, and 0.004 for SBayesR. However, differences between similarly performing methods were generally not statistically significant (due to moderately large standard errors), and thus caution should be exercised in their interpretation; for this reason, we did not perform secondary analyses to further assess differences between methods.

**Figure 5:**
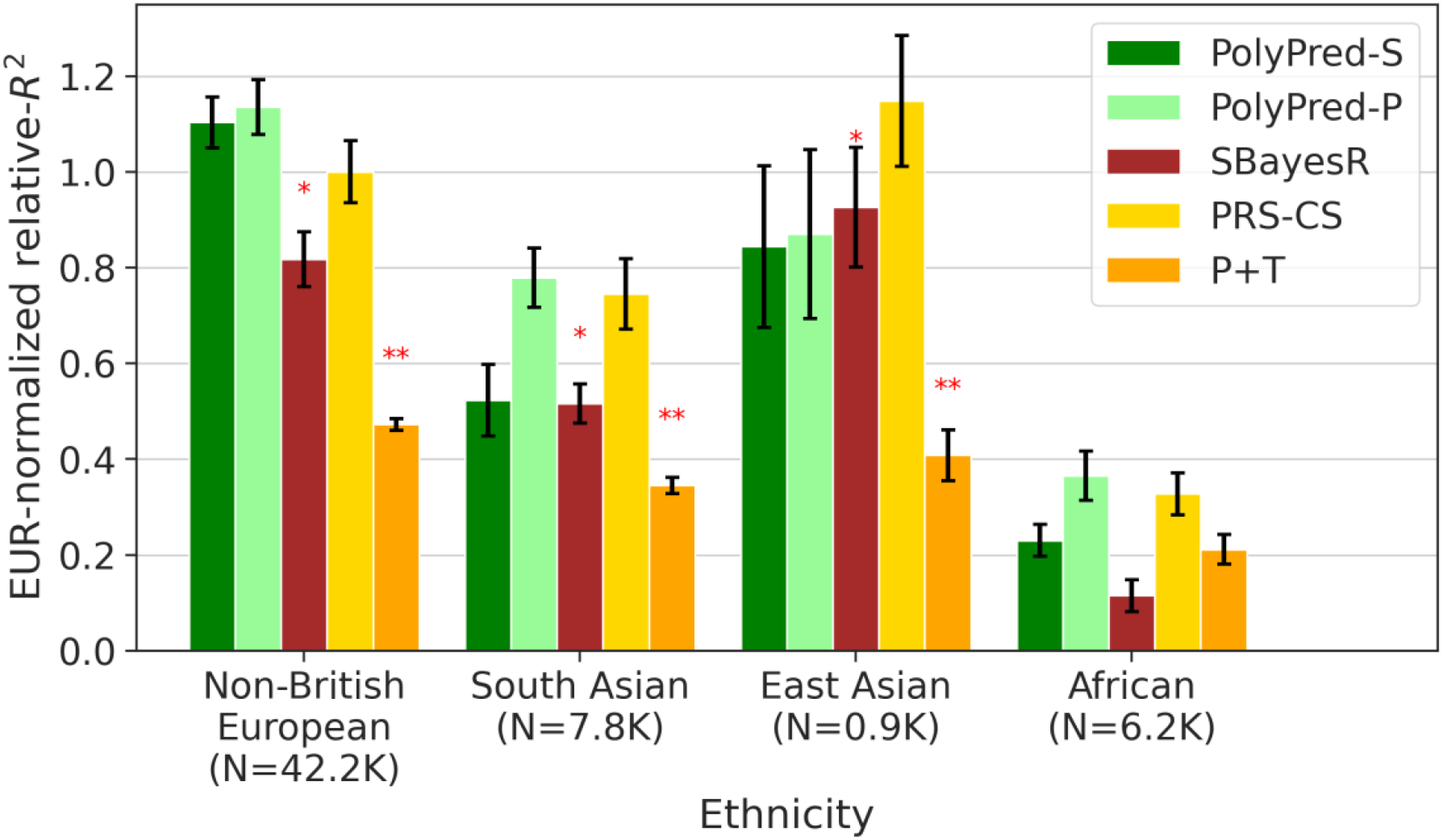
Cross-population PRS results for real UK Biobank traits, using summary statistics from a meta-analysis of many cohorts. We report average prediction accuracy (relative-*R*^2^; see main text), meta-analyzed across 4 well-powered, approximately independent traits, for PRS trained in European Network for Genetic and Genomic Epidemiology (ENGAGE) samples (average *N*=61,365) and applied to 4 UK Biobank populations. Target population sample sizes are indicated in parentheses; PolyPred and its summary statistic-based analogues used 500 additional training samples from each target population to estimate mixing weights. Asterisks above each bar denote statistical significance of the difference vs. PRS-CS, with red asterisks denoting a disadvantage (*P<0.05; **P<0.001). Errors bars denote standard errors. Numerical results, results for all 4 traits analyzed, absolute prediction accuracies (*R*^2^), and P-values of relative improvements vs. PRS-CS are reported in Supplementary Table 5 and Supplementary Table 8.

We conclude that PolyPred-P can increase cross-population polygenic prediction accuracy vs. published methods when analyzing summary statistics from a meta-analysis of many cohorts.

### Analysis of Biobank Japan and Uganda-APCDR cohorts

We applied PolyPred and its summary statistic-based analogues to predict 23 diseases and complex traits in Biobank Japan^41^ and 7 complex traits in Uganda-APCDR, an African-ancestry cohort^42,43^ (Methods, Supplementary Table 3). We performed these experiments to avoid training effect sizes and testing predictions in the same cohort, which may produce inflated prediction accuracies^33,72–74^. We again used UK Biobank British training data (average *N*=325K) to estimate SNP effect sizes, and used 500 individuals from the target population to estimate mixing weights. We evaluated prediction accuracy using individuals from each of the 2 target cohorts that were not included in the training data, and were unrelated to the training individuals and to each other: 5K Biobank Japan individuals and 1.3K Uganda-APCDR individuals. We again compared PolyPred and its summary statistic-based analogues to P+T, BOLT-LMM, SBayesR, and PRS-CS. We meta-analyzed relative-*R*^2^ across the same 7 well-powered, independent complex traits used in the UK Biobank analyses (Supplementary Table 3).

Results meta-analyzed across traits are reported in Figure 6, Supplementary Table 5, and Supplementary Table 9, and results for each trait are reported in Supplementary Table 5 and Supplementary Table 9. Among the published methods, we again observed that BOLT-LMM attained the highest prediction accuracy in each target population (although differences between BOLT-LMM, SBayesR, and PRS-CS were not statistically significant), and that P+T was substantially less accurate than the other methods, suffering relative losses of 42-61% vs. BOLT-LMM.

**Figure 6:**
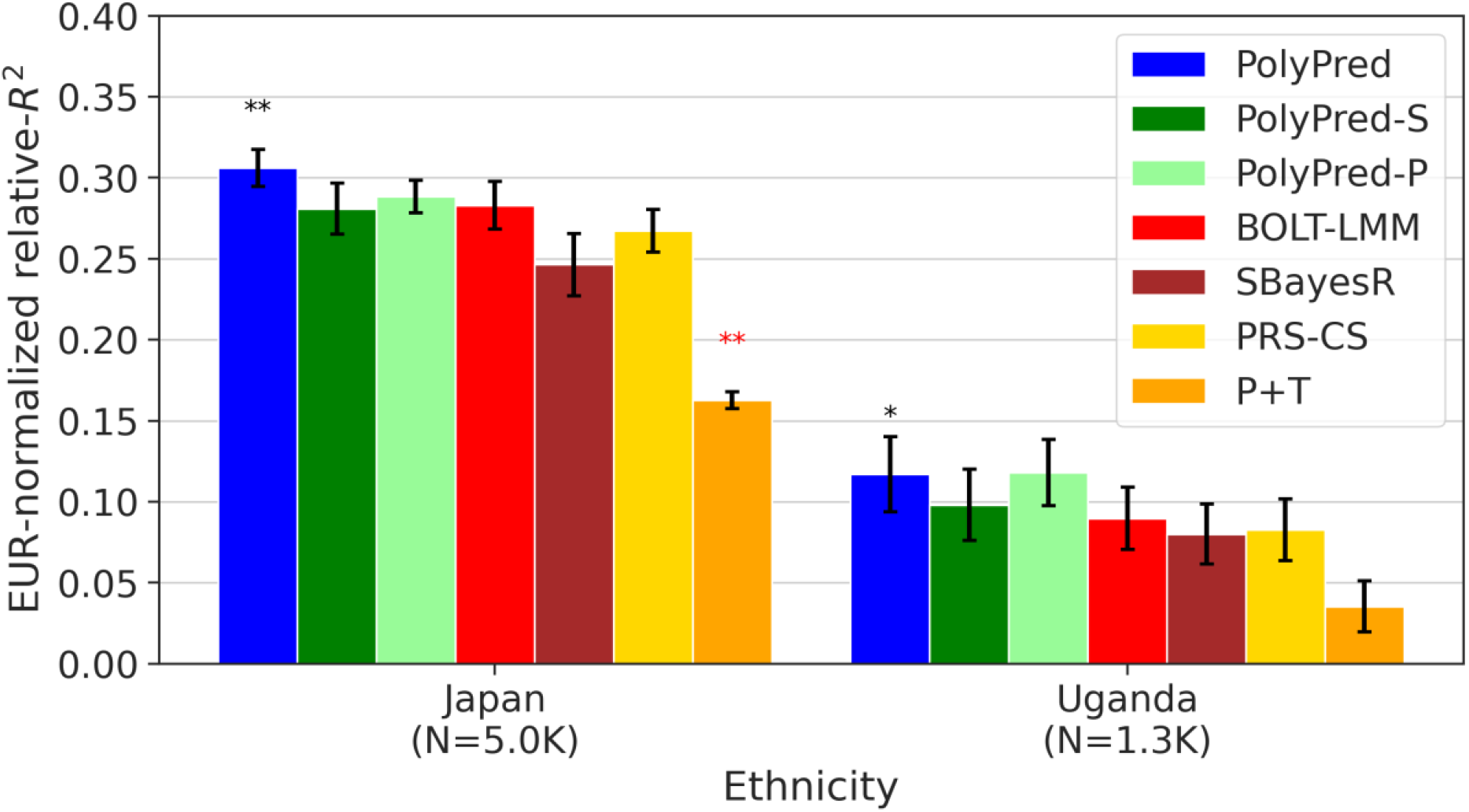
Cross-population PRS results for Biobank Japan and Uganda-APCDR traits. We report average prediction accuracy (relative-*R*^2^; see main text), meta-analyzed across 7 well-powered, independent traits, for PRS trained in UK Biobank British samples (average *N*=325K) and applied to Biobank Japan and Uganda-APCDR target populations. Target population sample sizes are indicated in parentheses; PolyPred and its summary statistic-based analogues used 500 additional training samples from each target population to estimate mixing weights. Asterisks above each bar denote statistical significance of the difference vs. BOLT-LMM, with black asterisks denoting an advantage and red asterisks denoting a disadvantage (*P<0.05; **P<0.001). Errors bars denote standard errors. Numerical results, results for all 23 traits analyzed, absolute prediction accuracies (*R*^2^), and P-values of relative improvements vs. BOLT-LMM are reported in Supplementary Table 9.

Among all 7 methods, PolyPred attained the highest prediction accuracy in Biobank Japan, and PolyPred-P attained the highest prediction accuracy in Uganda-APCDR (although the difference between PolyPred and PolyPred-P in Uganda-APCDR was not statistically significant). Improvements of PolyPred vs. BOLT-LMM in average relative-*R*^2^ were equal to +13% in Biobank Japan (*P*=2×10^−6^) and +22% in Uganda-APCDR (*P*=0.26), similar to our UK Biobank results above. We observed similar improvements for PolyPred-S vs. SBayesR and PolyPred-P vs. PRS-CS (both of which were statistically significant in Biobank Japan). Prediction accuracy (and hence relative-*R*^2^) for each method was much smaller in Biobank Japan and Uganda-APCDR (e.g. 0.32 and 0.11 for PolyPred; Figure 6) than in UK Biobank East Asians and UK Biobank Africans (0.62 and 0.34; Figure 4), likely due to higher SNP-heritabilities in the UK Biobank (see below). We also applied PolyPred+ and its summary statistic-based analogues to Biobank Japan, incorporating Biobank Japan training data (in addition to UK Biobank British training data) to estimate effect sizes (average *N*=124K, distinct from and unrelated to the 5K target individuals), with the caveat that this analysis involved training and testing in the same cohort (Methods). PolyPred+ attained increased prediction accuracy, with a further +23% improvement vs. PolyPred (*P*=0.0004), with similar results for PolyPred-S+ and PolyPred-P+ (although the improvement of PolyPred-P+ vs. PRS-CS was not statistically significant due to large standard errors) (Supplementary Tables 5,9).

We performed additional experiments to investigate the above result of decreased prediction accuracy in Biobank Japan vs. UK Biobank East Asians (of predictors trained using UK Biobank British training data). We compared BOLT-LMM trained using a reduced set of *N*=124K UK Biobank British training samples and applied to UK Biobank non-British Europeans vs. BOLT-LMM trained using the *N*=124K Biobank Japan training samples and applied to the Biobank Japan target samples. The prediction *R*^2^ of BOLT-LMM in UK Biobank non-British Europeans was +108% larger than in Biobank Japan, consistent with the +104% increase expected from theory^75,76^ based on the +67% higher SNP-heritabilities in UK Biobank (Supplementary Table 10, Supplementary Note). This suggests that differences in SNP-heritability due to ancestry differences (e.g. SNP ascertainment^67^, sample ascertainment, and/or ancestry-specific architectures^30^) or due to cohort differences (e.g. differences in phenotype definitions^13^, different recruiting strategies^13^, or assay artifacts) may explain most of the differences in prediction accuracies observed between the UK Biobank and Biobank Japan. Further experiments and interpretation are provided in the Supplementary Note.

We performed 6 secondary analyses. First, we assessed the calibration of each method by computing regression slopes (see above), which are reported in Supplementary Table 9. Similar to our above analyses of non-European UK Biobank target populations, PolyPred and its summary statistic-based analogues were the only approximately well-calibrated methods, as expected due to their extra training step to estimate mixing weights in the target population. We restricted the remaining secondary analyses to PolyPred (as PolyPred-S and PolyPred-P are analogous to PolyPred with respect to these analyses). Second, we evaluated a modification of PolyPred that estimates mixing weights using 500 UK Biobank individuals from the genetically closest target population (UK Biobank East Asians for Biobank Japan, UK Biobank Africans for Uganda-APCDR) instead of 500 individuals from the target cohort. The differences between the original and modified versions of PolyPred were small and not statistically significant (Supplementary Table 9), indicating that PolyPred mixing weights can be estimated using 500 individuals from any cohort with the same continental ancestry as the target population. Third, we evaluated modified versions of PolyPred that specify fixed mixing weights instead of estimating mixing weights in the target populations. We considered mixing weights for PolyFun-pred/BOLT-LMM equal to 0%/100%, 25%/75%, 50%/50%, 75%/25%, and 100%/0%. The 25%/75% and 50%/50% methods performed very similarly to PolyPred, with no statistically significant differences (Supplementary Table 9). Fourth, we reduced the number of training samples from the target population used to estimate mixing weights (*N*_mix_) from 500 to 100. PolyPred suffered slightly reduced accuracy but remained the most accurate method, with the improvement vs. BOLT-LMM in Biobank Japan remaining statistically significant (Supplementary Table 9). Fifth, we computed standard errors of relative-*R*^2^ using a jackknife over individuals^61^ (instead of a genomic block-jackknife over SNPs). We obtained standard errors that were almost identical to those obtained using a genomic block-jackknife (unlike the above results for UK Biobank), suggesting that Biobank Japan may be more heterogeneous across samples, possibly due to its hospital-based recruitment (Supplementary Table 9). Finally, we meta-analyzed the results of each method across three independent diseases in Biobank Japan: type 2 diabetes, asthma, and all autoimmune disease. Similar to our UK Biobank analyses above, PolyPred attained the highest prediction accuracy in each disease, though relative improvements were not statistically significant due to lower power (Supplementary Table 9).

We conclude that PolyPred and its summary statistic-based analogues substantially increase cross-population polygenic prediction accuracy vs. published methods when applied to target cohorts different from the training cohort.

### Analysis of UK Biobank East Asians using UK Biobank British and Biobank Japan training data

We applied PolyPred+ and its summary statistic-based analogues to predict 23 diseases and complex traits in UK Biobank East Asians using UK Biobank British and Biobank Japan training data (Supplementary Table 3). We performed this experiment to explore the special case where non-European training data is available in large sample size from a population that is genetically similar to the target population, in a cohort that is distinct from the target cohort; as such, this experiment is a particular strength of this study, relative to previous studies that considered only European training data or analyzed non-European training data from the same cohort as the target cohort^11,13–17^. We note that this experiment is still imperfect in that the European training data and non-European target data are from the same cohort (UK Biobank); however, we believe that cohort effects (if present) would deflate rather than inflate the relative improvement of PolyPred+ vs. other methods, since they would confer an advantage to the European training data but not the non-European training data. We used UK Biobank British training data (average *N*=325K) and Biobank Japan training data (average *N*=124K) to estimate SNP effect sizes. We again used 500 individuals from the target population to estimate mixing weights, and evaluated prediction accuracy using 900 UK Biobank East Asians that were not included in the training data, and were unrelated to the training individuals and to each other. We compared PolyPred, PolyPred+, and their summary statistic-based analogues to P+T, BOLT-LMM, SBayesR, and PRS-CS. When training SBayesR using Biobank Japan training data, we computed in-sample LD based on *N*=50K Biobank Japan individuals, following the recommendations of the authors of SBayesR^38^ (analogous to the LD matrices of European training data provided by the authors of SBayesR) (Methods). When training PRS-CS using Biobank Japan training data, we used East Asian LD matrices based on *N*=2,181 UK Biobank East-Asian individuals, provided by the authors or PRS-CS (Methods). We meta-analyzed relative-*R*^2^ across the same 7 well-powered, independent complex traits used in the previous analyses (Supplementary Table 3).

Results meta-analyzed across traits are reported in Figure 7 and Supplementary Tables 4-6, and results for each trait are reported in Supplementary Tables 4-6. PolyPred+ attained the highest prediction accuracy, with a +24% improvement vs. BOLT-LMM (*P*=0.0009) and a +12% improvement vs. PolyPred (*P*=0.0014). This implies that incorporating non-European training data can provide a substantial advantage, if it is available in large sample size. Results for PolyPred-S+ (vs. SBayesR and PolyPred-S) and PolyPred-P+ (vs. PRS-CS and PolyPred-P) were similar. We emphasize that the +12% improvement for PolyPred+ vs. PolyPred should be viewed as a lower bound on the improvement that would be obtained in settings without cohort effects that may confer an advantage to the European training data.

**Figure 7:**
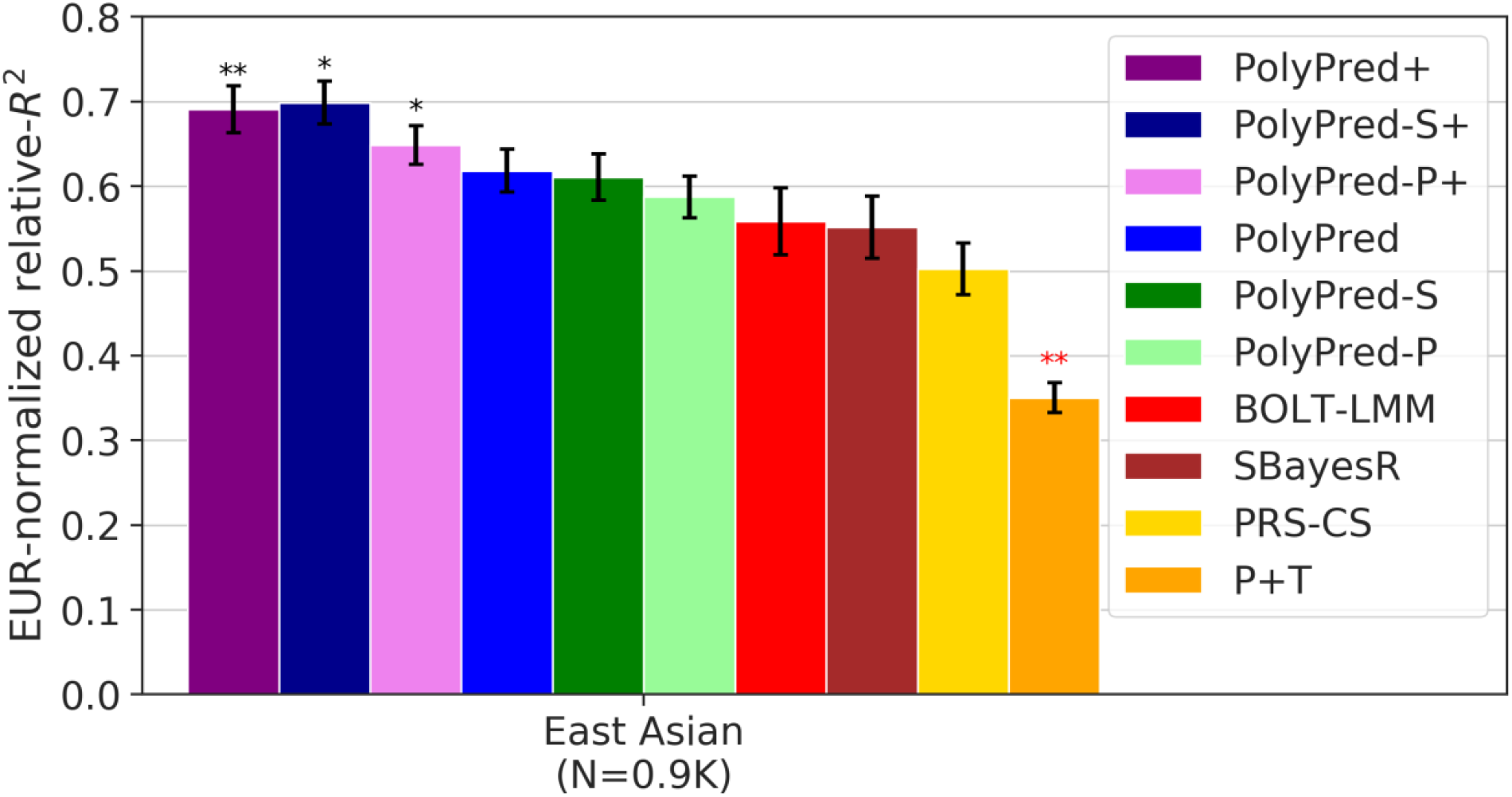
Cross-population PRS results for UK Biobank East Asians when incorporating both European and non-European training data. We report average prediction accuracy (relative-*R*^2^; see main text), meta-analyzed across 7 well-powered, independent traits, for PRS trained in UK Biobank British (average *N*=325K) and Biobank Japan samples (average *N*=124K; used by PolyPred+ and its summary statistic-based analogues only) and applied to UK Biobank East Asians. The target population sample size is indicated in parentheses; PolyPred, PolyPred+, and their summary statistic-based analogues used 500 additional training samples from the target population to estimate mixing weights. Asterisks above each bar denote statistical significance of the difference vs. BOLT-LMM, with black asterisks denoting an advantage and red asterisks denoting a disadvantage (*P<0.05; **P<0.001). Errors bars denote standard errors. Numerical results, results for all 23 traits analyzed, absolute prediction accuracies (*R*^2^), and P-values of relative improvements vs. BOLT-LMM are reported in Supplementary Tables 4-6.

We performed 6 secondary analyses. We restricted these secondary analyses to PolyPred+ (as PolyPred-S+ and PolyPred-P+ are analogous to PolyPred+ with respect to these analyses). First, we verified that PolyPred+ using European and East Asian training data does not outperform PolyPred in UK Biobank populations other than East Asians; differences between PolyPred+ and PolyPred were very small and not statistically significant (Supplementary Table 6). Second, we verified that PolyPred+ was well-calibrated (Supplementary Table 4; results for other methods are described above), as expected due to its extra training step to estimate mixing weights in the target population. Third, we evaluated a modified version of PolyPred+ that estimates mixing weights using 500 Biobank Japan individuals instead of 500 UK Biobank East Asians. The modified version of PolyPred+ was far less accurate than the original version (52% lower relative-*R*^2^; Supplementary Table 6). The mixing weights estimated in Biobank Japan assign much higher weight to the Biobank Japan training data (Supplementary Table 6), perhaps due to cohort effects; thus, it may be important to estimate PolyPred+ mixing weights using the target cohort (as opposed to the training cohort) if cohort effects are present. Fourth, we reduced the number of training samples from the target population used to estimate mixing weights (*N*_mix_) from 500 to 100. PolyPred+ suffered slightly reduced accuracy, though the difference was not statistically significant (Supplementary Table 6). Fifth, we evaluated a prediction method using only the *N*=124K Biobank Japan individuals to train effect sizes (BOLT-LMM-BBJ). BOLT-LMM-BBJ substantially underperformed methods that use UK Biobank British training data (−27% vs. BOLT-LMM, −34% vs. PolyPred, −41% vs. PolyPred+; Supplementary Table 4). Finally, we computed standard errors of relative-*R*^2^ using a jackknife over individuals^61^ (instead of a genomic block-jackknife over SNPs). Standard errors computed using a jackknife over individuals were smaller, increasing the statistical significance of relative improvements of PolyPred+ vs. other methods (Supplementary Table 6).

We conclude that PolyPred+ and its summary statistic-based analogues further increase cross-population prediction accuracy in the special case where non-European training data from the target population (or a closely related population) is available in large sample size. We emphasize that efforts to assess the benefit of incorporating non-European training data should analyze non-European training data from a cohort that is distinct from the target cohort, otherwise results may be inflated due to cohort effects.

## Discussion

We have introduced PolyPred, which improves cross-population polygenic risk prediction by incorporating causal effects in addition to tagging effects, addressing cross-population LD differences. Across seven well-powered independent traits, PolyPred significantly increased prediction accuracy over BOLT-LMM by 32% in UK Biobank Africans and by 13% in Biobank Japan (with similar results vs. SBayesR and PRS-CS). In the special case where a large training sample is available in the non-European target population (or a closely related population), we have introduced PolyPred+, which further incorporate the non-European training data, addressing MAF differences and causal effect size differences. PolyPred+ significantly increased prediction accuracy in UK Biobank East Asians over BOLT-LMM by 24% (and over PolyPred by 12%). PolyPred and PolyPred+ require individual-level training data (for their BOLT-LMM component), but we have also introduced summary statistic-based analogues of PolyPred and PolyPred+ in cases where individual-level training data is not available; specific recommendations are provided in Figure 2 (also see Table 2). We previously demonstrated that linearly combining PRS from European and non-European training samples improves cross-population prediction accuracy^7^. However, these previous results did not incorporate causal effects and used P+T, which is highly inaccurate despite its widespread use^11,13–18,23,31,52–56^, as PolyPred obtained up to 164% greater accuracy than P+T. In conclusion, PolyPred and its summary statistic-based analogues substantially improve cross-population polygenic prediction accuracy, ameliorating health disparities^13^. We have publicly released the PRS coefficients for all SNPs and traits analyzed under all evaluated methods (see URLs).

Although we substantially improved cross-population PRS accuracy over the state of the art, prediction accuracy in non-Europeans is still substantially lower compared to Europeans, even within the UK Biobank. There are two reasons for the remaining accuracy gap. First, European sample sizes are still limited, which limits the ability of PolyFun-pred to estimate causal rather than tagging effects (mathematical theory guarantees perfect estimation of causal effect sizes in European cohorts under an infinite sample size if model assumptions hold^77^). Second, non-European sample sizes are limited, which limits the ability of BOLT-LMM applied to non-European samples to estimate tagging effects. Even with an infinite European training sample, which allows estimating causal effects perfectly (thus addressing LD differences), prediction accuracy could still be higher for Europeans vs. non-Europeans due to cross-population genetic correlations less than 1^13,30,78,79^ and different allele frequencies (including population-specific SNPs) (Methods). (We note that cross-population genetic correlations less than 1 could potentially be explained by GxE interactions^80^, e.g. if G and GxE effects are shared across ancestries but the (average) value of E differs across ancestries^30^. However, if E is unmodeled, it is difficult to distinguish this scenario from the scenario of different G effects for other reasons.) Hence our theory and results confirm that larger non-European GWAS are the best way to further improve PRS accuracy in non-European populations^9,10,12,13,21^.

One of the main conclusions of our work is that leveraging training data from different ancestry groups (e.g. different continental ancestries) improves PRS in diverse populations. However, we recommend against using training data consisting of a traditional fixed-effect meta-analysis of GWAS data from different ancestry groups, for two reasons: (i) fixed-effect meta-analysis implies that European training samples and training samples from the non-European target population would receive equal weight, whereas our work shows that the latter should receive higher weight in order to maximize PRS accuracy; and (ii) it may be challenging to construct an LD reference panel whose ancestry matches the ancestry of the meta-analysis of different ancestry groups. When possible, it would be preferable to separately incorporate European training data and training data from the non-European target population, with appropriate LD reference panels. Although there is no single optimal way to choose a training cohort, training sample size should be a primary consideration, as it is a critical factor impacting PRS accuracy.

Our results corroborate previous results that predictions within the UK Biobank are often more accurate than off-cohort predictions to the same target ancestry^72–74^. This raises the question of whether the higher within-UK Biobank prediction accuracy is inflated by cohort effects. Our analysis suggests that within-UK Biobank prediction accuracy is not inflated, because most of the off-cohort loss of accuracy is driven by heritability differences. These heritability differences could be driven by between-cohort factors such as differences in phenotype definitions^13^, different recruiting strategies^13^, or assay artifacts. Our results are consistent with recent results showing almost no loss of accuracy when applying PRS based on UK Biobank training data to other European-ancestry cohorts^38^. Importantly, our results suggest that factors that inflate within-cohort PRS accuracy^81^ (such as cohort-specific GxE, cohort-specific indirect effects^82^, cohort-specific population structure, or cohort-specific assortative mating) are unlikely explanations for the observed accuracy differences between the UK Biobank and Biobank Japan.

Our work has several limitations, providing opportunities for future work. First, we did not evaluate a setting where the British training data, the non-British training data, and the target population are sampled from three different cohorts. However, we hypothesize that the relative improvement of PolyPred+ over PolyPred when applied to UK Biobank East Asians reflects a lower bound on the improvement in relative-*R*^2^ that would have been obtained in such an experiment. Second, PolyPred and its summary statistic-based analogues are slower than alternative PRS methods, requiring over 1,000 hours of computation time for training, vs. less than 100 hours for BOLT-LMM. This is dominated by the PolyFun-pred component, which is computationally intensive because (i) PolyFun-pred performs fine-mapping, which is a more computationally intensive task than other approaches to computing PRS coefficients (e.g. computing posterior mean tagging effect sizes, as in SBayesR); and (ii) PolyFun-pred analyzes a large number of SNPs, e.g. 18 million SNPs in UK Biobank training data and 8.1 million SNPs in ENGAGE training data (vs. 1.2 million SNPs for SBayesR). We do not foresee the larger computation time for training as a major limitation in real-world settings, because training only needs to be performed once, can be parallelized, and provides genome-wide fine-mapping results of direct interest^35^. Third, PolyPred requires a large number of SNPs (e.g. 8.1 million SNPs in the ENGAGE analysis) to perform fine-mapping. In addition, these SNPs must be imputed in the target sample (whereas BOLT-LMM uses only HapMap 3 SNPs, which are typically well-imputed across most cohorts^83^), motivating the need for large cross-population imputation panels (PolyPred becomes far less accurate when using only HapMap 3 SNPs). We note that naively scaling BOLT-LMM, SBayesR, or PRS-CS to use such a large number of SNPs would be computationally infeasible. Fourth, our block-jackknife standard error estimates may be conservative, though they may be better suited for evaluating the sampling variance introduced by the training set (vs. individual-level jackknife, which assumes a fixed training set; see Methods). Fifth, our PRS do not capture effects from the HLA region, which explains a large proportion of the variance of several diseases and traits, owing to the very complex and long-range LD patterns in this region. Sixth, PolyPred requires a large European training sample to perform accurate fine-mapping (we recommend *N*>100K based on previous studies^35^). Seventh, PolyPred+ requires a large training sample that is closely related to the target population. However, it is not clear exactly how large this sample should be (we currently recommend *N*>50K), or how to quantify genetic similarity between the training and target populations (as LD differences between populations are driven by divergence rather than genetic drift^58^). Eighth, PolyPred ideally requires a small training sample from the target cohort to estimate mixing weights. Our results suggest that it is possible to improve cross-population PRS accuracy even without such a training sample, by linearly combining PolyFun-pred and BOLT using mixing weights of either 25%/75% or 50%/50%, respectively. However, we caution that PRS linearly combined using fixed mixing weights may not always be well-calibrated. Ninth, it may be preferable to construct a European and a non-European PRS jointly^24,25^, rather than linearly combining a European and a non-European PRS as performed in PolyPred+. Tenth, it may be possible to improve PRS accuracy for admixed individuals by using European effect sizes for European alleles and non-European effect sizes for non-European alleles^16,17^. Eleventh, cross-population prediction accuracy may be improved by identifying SNP sets other than HapMap 3 that yield better prediction accuracy across cohorts. Twelfth, prediction accuracy could in principle be improved if it were possible to decompose the PolyFun-pred and BOLT-LMM predictors into shared and non-shared components, to improve upon double counting of shared components vs. single counting of non-shared components (Supplementary Note). Thirteenth, prediction accuracy could potentially be improved by applying PolyFun-pred to non-European training data and incorporating this predictor (although existing non-European training samples are generally not large enough to reap the benefits of PolyFun-pred). Finally, PolyPred may be able to estimate causal effect sizes more accurately by using a cross-population fine-mapping method (instead of PolyFun-pred, which uses only European training data). Despite all these limitations, PolyPred and PolyPred+ and their summary statistic-based analogues provide a clear improvement for cross-population polygenic risk prediction.

## Methods

### PolyPred and its summary statistic-based analogues

All methods in this paper use a linear PRS, i.e., 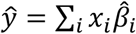, where *ŷ* is the PRS of an individual, *x*_*i*_ is the number of minor alleles of SNP *i* carried by that individual, and 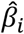is the estimated per-allele causal effect size of SNP *i*. The methods differ in the way they estimate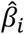.

PolyPred and PolyPred+ both combine the methods PolyFun-pred and BOLT-LMM; PolyPred-S and PolyPred-S+ both combine the methods PolyFun-pred and SBayesR; and PolyPred-P and PolyPred-P+ both combine the methods PolyFun-pred and PRS-CS. PolyFun-pred estimates 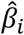 as the (approximate) posterior mean causal effect size of SNP *i*, as estimated by PolyFun + SuSiE^35^ based on European training data, using 187 functional annotations to specify prior causal probabilities (see below). BOLT-LMM (resp. SBayesR and PRS-CS) estimates tagging effects (Supplementary Note) of HapMap 3 SNPs by applying BOLT-LMM^36,37^ (resp. SBayesR^38^ and PRS-CS^39)^ to European training data. BOLT-LMM (resp. SBayesR) treats the effect of each SNP *i* as a random effect sampled from a mixture of two (resp. four) zero-mean normal distributions, whose variances and mixture weights are determined in a data-driven manner. PRS-CS treats the effect of each SNP *i* as a random effect sampled from a continuous shrinkage prior distribution.

PolyPred and its summary statistic-based analogues compute the effect size of each SNP *i* that is either in HapMap 3 or has a European MAF≥0.1% and INFO score ≥0.6 as a weighted combination of (1) its PolyFun-pred effect size based on European training data; and (2) its BOLT-LMM (resp. SBayesR and PRS-CS) effect size based on European training data:

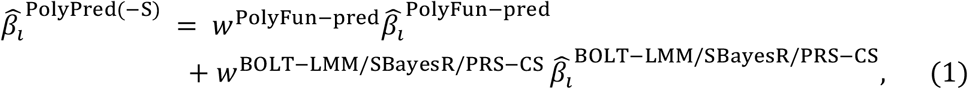

where 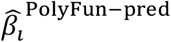 is the PolyFun-pred approximate posterior mean causal effect size of SNP *i* based on European training data, 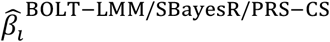 is the approximate posterior mean tagging effect size of SNP *i* based on European training data using the indicated method (setting the effects of SNPs not in HapMap 3 to zero), and *w*^PolyFun−pred^, *w*^BOLT−LMM/SBayesR/PRS−CS^ are mixing weights. PolyPred estimates the mixing weights via non-negative least squares estimation (i.e., least squares estimation constrained to produce to non-negative estimates) based on training individuals from the target cohort. Specifically, PolyPred (resp. PolyPred-S and PolyPred-P) estimates the mixing weights by computing the PRS corresponding to the PolyFun-pred effect sizes (given by 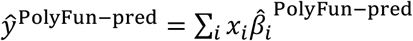) and the PRS corresponding to the BOLT-LMM (resp. SBayesR and PRS-CS) effect sizes (given by 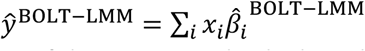), and then fitting the mixing weights by regressing the true phenotypes *y*_*i*_ of the training individuals in the target cohort on the PolyFun-pred and the BOLT-LMM (resp. SBayesR and PRS-CS) PRSs. The use of non-negative least squares estimation guarantees that the correlation of the predicted phenotype with the true phenotype is at least as large as the smallest correlation obtained by the constituent predictors.

PolyPred+ and its summary statistic-based analogues compute the effect size of each SNP *i* that is either in HapMap 3 or has a European MAF≥0.1% as a weighted combination of (1) its PolyFun-pred effect size based on European training data; (2) its BOLT-LMM (resp. SBayesR and PRS-CS) effect size based on European training data; and (3) its effect size as estimated by applying BOLT-LMM (resp. SBayesR and PRS-CS) to training data from the target population (or a closely related population):

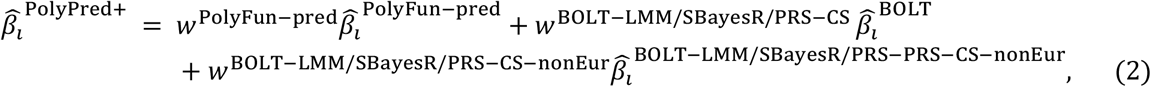

where 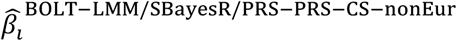 is the BOLT-LMM (resp. SBayesR or PRS-CS) approximate posterior mean tagging effect of SNP *i* based on training data from the non-European population (and set to zero for SNPs that are not in HapMap 3), and *w*^BOLT−LMM/SBayesR/PRS−CS−nonEur^ is the mixing weight of 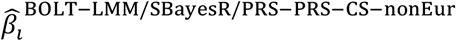. The mixing weights are estimated as in PolyPred. We note that the weighting used by PolyPred and its summary statistic-based analogues may be suboptimal if the correlations between PolyFun-pred effect sizes and BOLT-LMM effect sizes (resp. SBayesR and PRS-CS effect sizes) vary across the genome (Supplementary Note).

In practice, we apply PolyPred and its summary statistic-based analogues by linearly combining the PolyFun-pred PRS and the BOLT-LMM (or SBayesR or PRS-CS) PRS (rather than linearly combining the SNP effect sizes). The two procedures are almost mathematically identical, with the only difference being that a linear combination of PRSs can also accommodate an intercept, which explicitly bias-corrects the PRS to the target population.

We applied PolyFun-pred in the same way that we applied PolyFun + SuSiE in our previous work^35^. Briefly, we applied PolyFun-pred across 2,763 overlapping 3Mb loci (equally spaced starting at chromosome 1, position 0) spanning 18,212,157 European MAF>0.1% imputed SNPs with INFO score>0.6 (excluding the HLA and two other long-range LD regions)^35^, assuming 10 causal SNPs per locus. We used summary statistics computed by BOLT-LMM, based on up to *N*=337,491 unrelated British-ancestry UK Biobank individuals, and using summary LD information estimated directly from the target samples. Full details are provided in ref. ^35^. We note that the use of BOLT-LMM summary statistics is mathematically equivalent to regressing the target phenotypes on BOLT-LMM off-chromosome PRS prior to applying PolyFun + SuSiE^37^. We also note that the use of 3Mb loci guarantees that for each SNP, the estimation of its causal effect size takes into account virtually all relevant SNPs that may be in LD with that SNP (because LD in European populations rarely ranges beyond 1Mb^59^), allowing to disentangle its causal effect size from its tagging effect size.

In secondary analyses, we evaluated alternative versions of PolyFun-pred that assume a single causal SNP per locus (and hence do not require an LD reference panel^35^) or a non-functionally-informed version that specifies the same prior causal probability to all SNPs in each locus.

PRS methods that include non-common SNPs (MAF<5%) may be sensitive to MAF-dependent and LD-dependent architectures (e.g., low-MAF SNPs have a smaller average per-SNP heritability^44–46^). Previous PRS methods have largely alleviated this concern by discarding non-common SNPs instead of explicitly modeling their lower per-SNP heritability^33,38,39,61–63,72,73,84–86^. In contrast, PolyFun-pred accounts for MAF-dependent and LD-dependent architectures by specifying SNP-specific prior causal probabilities based on the baseline-LF model^45^ (Supplementary Table 11). In detail, PolyFun-pred used 187 overlapping functional annotations from the baseline-LF model (previously described in ref. ^35^), including 10 common MAF bins (MAF≥0.05); 10 low-frequency MAF bins (0.05>MAF≥0.001); 6 LD-related annotations for common SNPs (levels of LD, predicted allele age, recombination rate, nucleotide diversity, background selection statistic, CpG content); 5 LD-related annotations for low-frequency SNPs; 40 binary functional annotations for common SNPs; 7 continuous functional annotations for common SNPs; 40 binary functional annotations for low-frequency SNPs; 3 continuous functional annotations for low-frequency SNPs; and 66 annotations constructed via windows around other annotations^87^ (Supplementary Table 11). We did not include a base annotation that includes all SNPs, because such an annotation is linearly dependent on all the MAF bins when using the same set of SNPs to compute LD-scores and to estimate annotation coefficients^35^.

### Estimating relative-*R*^2^ and its standard error

We measured prediction accuracy for each trait via a measure that we call relative-*R*^2^, defined via the following computations:

1. Compute *R*^2^-PRS: the *R*^2^ obtained via a linear predictor that includes PRS, age, sex, age*sex (if the correlation with age was <0.95), UK Biobank assessment center (defined as a set of dummy binary variables), genotyping array, 10 principal components (computed separately for each ancestry; see below), and dilution factor (for biochemical traits only).
2. Compute *R*^2^-noPRS, defined like *R*^2^-PRS but omitting the PRS
3. Compute *R*^2^-PRS-BOLT-EUR, computed by applying BOLT-LMM to UK Biobank non-British Europeans as in step 1
4. Compute *R*^2^-noPRS-BOLT-EUR, computed by applying step 2 to non-British Europeans
5. Compute relative-*R*^2^ as (*R*^2^-PRS - *R*^2^-noPRS) / (*R*^2^-PRS-BOLT-EUR - *R*^2^-noPRS-BOLT-EUR) We note that relative improvement in relative-*R*^2^ is the same as relative improvement in absolute difference in *R*^2^, (i.e., in *R*^2^-PRS - *R*^2^-noPRS), because the denominator (*R*^2^-PRS-BOLT-EUR - *R*^2^-noPRS-EUR) can be regarded as a trait-specific scaling factor.

We computed standard errors via genomic block-jackknife, partitioning the genome into 200 equally-sized consecutive loci and omitting each one in turn. We similarly computed standard errors of differences in relative-*R*^2^ (e.g. vs. BOLT-LMM) via genomic block-jackknife, computing the difference after omitting each block in turn. In secondary analyses, we computed standard errors by applying jackknife over individuals from the target population. These analyses yielded much smaller standard errors in the UK Biobank, suggesting that genomic block-jackknife standard errors may be conservative, whereas individual-based jackknife estimates maty be anti-conservative. We emphasize that individual-based jackknife explicitly assume a fixed training set.

We meta-analyzed relative-*R*^2^ via an inverse-variance weighted average, using weights inversely proportional to the standard error of the *R*^2^ of BOLT-LMM in the target ancestry (as estimated via genomic block-jackknife). We estimated the standard error of the meta-analyzed relative-*R*^2^ as the square root of the weighted average of the trait-specific sampling variances, divided by the number of traits. We meta-analyzed the difference in relative-*R*^2^ vs. an alternative method (typically BOLT-LMM) in the same way. We computed p-values of differences in relative-*R*^2^ via a Wald test, based on the block-jackknife standard error estimates (for single traits) and based on the meta-analyzed standard errors (for the meta-analyzed results).

We computed ancestry-specific regression slopes by regressing true phenotypes on the PRS (including an intercept) in each respective population. We computed the standard errors of regression slopes via genomic block-jackknife, using 200 jackknife blocks.

We computed the statistical significance of the decrease in *R*^2^ in non-European vs. European target samples via a Wald test for the difference in *R*^2^, conservatively estimating the sampling variance of this difference as the sum of the sampling variances of the European *R*^2^ and the non-European *R*^2^ (this is a conservative estimate as long as the *R*^2^ estimates in Europeans and non-Europeans are not negatively correlated, which is extremely unlikely).

### Cohorts Analyzed

#### UK Biobank

The UK Biobank is a UK-based population cohort^40^. We used version 3 of the imputed genotypes, as described in our previous work^35^. We computed ancestry-specific PCs for UK Biobank Africans, UK Biobank East Asians, and UK Biobank South Asians via plink 1.9^88^, restricting to SNPs with ancestry-specific MAF>5%, missingness<10%, HWE p-value>10^−10^, and LD-pruned using the command --indep-pairwise 1000 50 0.05, and restricted to unrelated individuals (kinship coefficient <0.05) from the target ancestry with missingness <10%. We used the UK Biobank provided PCs for UK Biobank Europeans.

We defined the ‘autoimmune disease’ trait in the UK Biobank as a union of the following UK Biobank codes: 1154 (irritable bowel syndrome); 1222 (type 1 diabetes); 1224 (thyroid problem); 1225 (hyperthyroidism/thyrotoxicosis); 1226 (hypothyroidism/myxoedema); 1256 (acute infective polyneuritis/guillain-barre syndrome); 1260 (myasthenia gravis); 1261 (multiple sclerosis); 1313 (ankylosing spondylitis); 1372 (vasculitis); 1377 (polymyalgia); 1378 (wegners granulmatosis); 1381 (systemic lupus erythematosis/sle); 1382 (sjogren’s syndrome/sicca syndrome); 1384 (scleroderma/systemic sclerosis); 1437 (myasthenia gravis); 1453 (psoriasis); 1456 (malabsorption/coeliac disease); 1461 (inflammatory bowel disease); 1462 (Crohns disease); 1463 (ulcerative colitis); 1464 (rheumatoid arthritis); 1477 (psoriatic arthropathy); 1522 (grave’s disease); 1661 (vitiligo); 1667 (alopecia / hair loss).

##### European Network for Genetic and Genomic Epidemiology

European Network for Genetic and Genomic Epidemiology (ENGAGE) is a consortium comprised of 24 cohorts to study the impact of genetic variations on medical phenotypes (e.g., type 2 diabetes, BMI, lipid phenotypes, etc.) through GWAS^68^. The consortium has performed over 80,000 GWASs using genetic and phenotype samples from over 600,000 individuals, and made the GWAS summary statistics publicly available^68^.

We obtained ENGAGE GWAS summary statistics, representing fixed-effect meta-analyses from 22 studies of European ancestry, for 2 lipid phenotypes^69^ (triglyceride (N=56,267) and total cholesterol (N=58,327)), and 2 obesity-related phenotypes^70^ (BMI (N=80,938) and BMI-adjusted waist hip ratio (N=49,877)). In each ENGAGE study, up to 37.4 million autosomal variants were imputed using the 1000 Genomes Project (we used 8.1 million variants which were also imputed in the UK Biobank); phenotypes were adjusted for age, age squared, genotype principal components, and other study-/trait-specific covariates, and were inverse rank normalized; GWASs were performed for each sex separately and combined using fixed-effect meta-analysis; a single genomic control correction was performed for each study prior to a cross-study meta-analysis^69,70^.

#### Biobank Japan

BioBank Japan (BBJ) is a multi-institutional hospital-based biobank with DNA and serum samples from approximately 200,000 participants from 12 medical institutions in Japan^41^. The participants are mainly of Japanese ancestry and had been diagnosed with at least one of 47 diseases by physicians at the cooperating hospitals. Written informed consent was obtained from all the participants, as approved by the ethics committees of RIKEN Center for Integrative Medical Sciences and the Institute of Medical Sciences at the University of Tokyo.

We genotyped samples with either (i) the Illumina HumanOmniExpressExome BeadChip or (ii) a combination of the Illumina HumanOmniExpress and HumanExome BeadChips. We applied standard quality control criteria for both samples and variants as detailed elsewhere^89^. We then pre-phased genotypes with Eagle2^90^ and imputed dosages with Minimac3^91^ using 1000 Genomes project phase 3 (version 5) data (*N*=2,504) and Japanese whole-genome sequencing (WGS) data (*N*=1,037) as a reference^89^. We computed PCs using EIGENSOFT’s smartpca^92^.

For phenotypes, we retrieved clinical medical records from the participating hospitals through interviews and a standardized questionnaire. We used 23 diseases and complex traits in Biobank Japan which are also analyzed in UK Biobank (Supplementary Table 3). We normalized quantitative phenotypes via inverse-rank normal transformation as described elsewhere^93^. We defined the ‘autoimmune disease’ trait in Biobank Japan as a union of Graves’ disease and rheumatoid arthritis patients.

#### Uganda-APCDR

Uganda-APCDR is a population-based cohort from the General Population Cohort (GPC), Uganda. We retrieved genotype and phenotype data through the African Partnership for Chronic Disease Research (APCDR) initiative via the European Genome-Phenome Archive (EGA), using EGAD00010000965 to access genotype data. Phenotype data were accessed via sftp from EGA (reference: DD_PK_050716 gwas_phenotypes_28Oct14.txt). The participants are from nine ethno-linguistic groups in sub-Saharan Africa and had been recruited from the study area located in southwestern Uganda in Kyamulibwa subcounty of Kalungu district, approximately 120 km from Entebbe town. These ethno-linguistic groups have diverse population structure with varying degrees of admixture between Eurasian and East African Nilo-Saharan ancestries, which has been extensively characterized elsewhere^94^. The detailed cohort demographics, sample collection, and processing were described previously^42,43^.

Briefly, the samples were genotyped using the Illumina HumanOmni 2.5M BeadChip at the Wellcome Trust Sanger Institute. We used the Ricopili pipeline to conduct pre-imputation QC and perform phasing and imputation^95^. Briefly, we phased the data using Eagle 2.3.5^90^ and imputed variants using minimac3^91^ in chunks ≥3Mb. The 1000 Genomes project phase 3 haplotypes^59^ were used as the reference panel for phasing and imputation.

As described previously, phenotypes were collected using a standard individual questionnaire, blood samples (laboratory tests), and biophysical measurements (height, weight, waist and hip circumferences and blood pressure)^42^. We normalized quantitative phenotypes via inverse-rank normal transformation.

### UK Biobank Simulations

We simulated data based on real genotypes of UK Biobank individuals, using 250,963 MAF≥0.1% SNPs with INFO score≥0.6 on chromosome 22 (including short indels). To simulate data, we first computed the variance of per-standardized-genotype effect *η*_*i*_ for every SNP *i* with annotations ***a***_*i*_ using the baseline-LF (version 2.2.UKB) model, 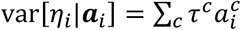, where *c* are annotations and *τ*^*c*^ estimates are taken from a fixed-effects meta-analysis of 16 well-powered genetically uncorrelated (|*r*_*g*_|<0.2) UK Biobank traits (age of menarche, BMI, balding, bone mineral density, eosinophil count, FEV1/FVC ratio, forced vital capacity, hair color, height, platelet count, red blood cell distribution width, red blood cell count, systolic blood pressure, tanning, waist-hip ratio adjusted for BMI, white blood count), scaled such that *∑*_*i*_var[*η*_*i*_|***a***_*i*_] is the same across all traits (as detailed in ref.^35^). Each SNP was specified to be causal with probability proportional to var[*η*_*i*_|***a***_*i*_], such that the average causal probability was equal to the desired proportion of causal SNPs (0.1% or 0.3% in most simulations).

We generated ancestry-specific effect sizes as follows. First, we generated a British per-allele causal effect size for each SNP *i* via 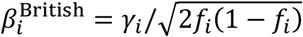, where *γ*_*i*_ ∼ 𝒩(0, *h*^2^/*m*), *m* is the number of causal SNPs, and *f*_*i*_ is the maximal MAF of SNP *i* among British, non-British European, South Asian, East Asian, or African UK Biobank individuals. Afterwards, for each of the main UK Biobank non-European ancestries (South Asian, East Asian, and African) *a* we generated an ancestry-specific per-allele effect size 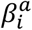 via 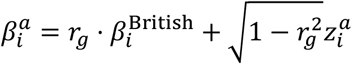, where *r*_g_ is the cross-population genetic correlation (set to 0.8 by default, following previous works^30,78,79^), and 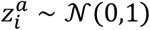. The use of *f*_*i*_ bounds the per-allele causal effect sizes by the MAF of the ancestry in which the SNP is most common, which guarantees that SNPs that are infrequent in Europeans but are common in other ancestries do not explain a very large proportion of heritability.

After generating ancestry-specific per-allele causal effect sizes, we generated a phenotype *y* for every UK Biobank individual in each ancestry *a* via 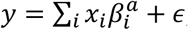, where *x*_*i*_ is the number of minor alleles of SNP *i* carried by that individuals, 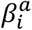 is the ancestry-specific per-allele causal effect size of SNP *i*, and *ϵ* ∼ 𝒩(0,1 − *h*^2^) is the environmental variance of the generated trait. We generated phenotypes based on dosage data from imputed genotypes, using Plink 2.0^96,97^. We used self-reported ancestry based on UK Biobank data field 21000 (Ethnic background). We considered Irish-ancestry as a non-British European ancestry.

We trained all methods using 337,491 unrelated British-ancestry individuals^40^, and we estimated the mixing weights of PolyPred and its summary statistic-based analogues using up to 1000 additional individuals from each of the four non-British ancestries (*N*_mix_≤1000). We computed summary statistics by applying linear regression via Plink 2.0. We did not evaluate PolyPred+ in the simulations because of the relatively small sample sizes of the UK Biobank non-European populations.

We evaluated prediction accuracy via *R*^2^, using held-out individuals that were not included in the training sets, using 42K non-British Europeans, 7.7K South Asians, 0.9K East Asians, and 6.2K Africans. We computed PRSs by applying plink 2.0 with the --score command, using imputed dosage data (rather than hard-called SNP values). We computed standard errors via a jackknife over simulations.

We trained BOLT-LMM by applying BOLT-LMM v2.3.4 to plink files of HapMap 3 SNPs (hard-coded from imputed dosages), using the same covariates specified in the “Estimating relative-*R*^2^ and its standard error” Methods subsection, and specifying the flag –predBetasFile to report PRS coefficients.

We trained SBayesR using summary statistics from the infinitesimal version of BOLT-LMM (BOLT-LMM-inf^36^), which yielded far superior accuracy vs. using summary statistics from the non-infinitesimal version of BOLT-LMM (BOLT-LMM) (results not shown), possibly indicating that the non-infinitesimal version of BOLT-LMM violates some of the underlying assumptions behind the SBayesR model. We ran SBayesR using 10,000 iterations, 4,000 burn-in iterations, using values from 10% of the iterations to compute posterior means, using the HapMap 3 LD files published the SBayesR authors^51^. We attempted to run SBayesR using a mixture of four distributions (using *π* = [0.95,0.02,0.02,0.01] and *γ* = [0,0.01,0.1,1]). In case SBayesR failed with these parameters, we iteratively shrank the last entry in the vector *γ* by 50% until it was smaller than 10^−6^, at which point we removed the last mixture component and redefined *π* such that the first entry was equal to 0.95 and all other entries had the same value such that all values sum to 1.0.

We trained PRS-CS using summary statistics from BOLT-LMM-inf (as in SBayesR) with the parameters *a*=1, *b*=0.5, thin=5, n_iter=10000, n_burnin=500, and without specifying the value of phi (corresponding to PRS-CS-auto). We used the UK Biobank LD reference panels made publicly available by the authors of PRS-CS (see URLs). We did not compute additional LD reference panels because the PRS-CS software does not provide this capability.

We trained P+T by applying plink with the command –clump-r2 0.5 –clump-kb 250 with various values of –clump-p1 (following ref.^13^), and using 10,000 randomly selected unrelated UK Biobank British individuals to compute LD. We estimated LD using 10,000 individuals to balance between runtime and accuracy (noting that P+T is relatively insensitive to the LD reference panel size compared to the other methods evaluated in this manuscript). We used summary statistics based on BOLT-LMM, using marginal effect sizes derived from reported 𝒳^2^ values (i.e., the square root of the 𝒳^2^ divided by the square root of the BOLT-LMM effective sample size^35^ and multiplied by the sign of the effect size estimated by the infinitesimal version of BOLT-LMM), because the non-infinitesimal version of BOLT-LMM does not estimate effect sizes. We used the best value of –clump-p1 (out of the evaluated values 10^−2^, 10^−3^, 10^−4^, 10^−6^, 5×10^−8^) based on the target sample phenotypes, which leads to anti-conservative prediction accuracy estimates for P+T.

When modifying the training sample size, we kept the LD reference panel sample size fixed to alleviate computational costs.

We used slightly different LD reference panels for PolyFun-pred, SBayesR, and PRS-CS, because (i) they use different algorithms to impose sparsity on LD matrices, and different file formats to store them; and (ii) we assume that naively running SBayesR or PRS-CS using summary LD from the 18 million SNPs used by PolyFun-pred would be computationally infeasible, based on information provided in the manuscripts describing these methods^38,39^. As noted above, increasing the number of SNPs analyzed by SBayesR from 1.2 million to 2.8 million does not improve the prediction accuracy of SBayesR (Supplementary Table 4).

### Analysis of UK Biobank, Biobank Japan and Uganda-APCDR cohorts

We performed three sets of analyses: (i) Analysis of 4 UK Biobank populations using UK Biobank British training data; (ii) Analysis of Biobank Japan and Uganda-APCDR cohorts; and (iii) Analysis of UK Biobank East Asians using UK Biobank British and Biobank Japan training data. In all analysis sets, we evaluated PRSs generated by training all methods using unrelated UK Biobank British-ancestry individuals. In a subset of analysis set (ii) and in analysis set (iii) we additionally evaluated PRSs generated by training BOLT-LMM-BBJ (BOLT-LMM trained on Biobank Japan individuals). The details below pertain to all three analysis sets unless specified otherwise.

We selected the 7 traits to meta-analyze by first restricting the set of 49 traits analyzed in ref.^35^ to traits that are available in Biobank Japan and Uganda-APCDR and are well-powered across multiple ancestries, having *h*^2^>0.05 in UK Biobank non-British Europeans, in UK Biobank South Asians, and in UK Biobank Africans (see below for details on ancestry-specific heritability estimation). We then iteratively greedily selected ranked traits according to their heritability in UK Biobank non-British Europeans (estimated as in ref. ^35^), such that no selected trait had |*r*_g_|<0.3 with a previously selected trait.

We computed ancestry-specific SNP heritabilities in each UK Biobank ancestry by applying GCTA^64^ to unrelated sets of individuals using hard-called HapMap 3 SNPs (using a random set of 10,000 individuals for non-British Europeans to facilitate the computations). We did not use more advanced methods^98^ because of the relatively small sample sizes. We meta-analyzed ancestry-specific SNP heritabilities by averaging the estimated heritabilities, and we estimated the meta-analyzed standard error via the square root of the average sampling variance, divided by the square root of the number of traits.

We trained all PRS methods on UK Biobank unrelated British-ancestry individuals (average *N*=325) as described in the Methods subsection “UK Biobank simulations”, but using summary statistics generated by BOLT-LMM when applied to UK Biobank British-ancestry individuals, as described in our previous work^35^. We trained P+T separately for each non-UK Biobank cohort by restricting the set of SNPs considered to the set of SNPs available in the target cohort. We computed the contribution of PolyFun-pred (resp. BOLT-LMM) towards PolyPred via the ratio of the mixing weight of PolyFun-pred (resp. BOLT-LMM) to the sum of the mixing weights of PolyPred and of BOLT-LMM.

In analysis sets (i) and (iii), we computed a PRS for each UK Biobank individual using imputed dosage data as described in the “UK Biobank Simulations”. In analysis set (ii), we computed a PRS for each individual in Biobank Japan and in Uganda-APCDR using imputed dosage data and PRS coefficients from UK Biobank Europeans using Plink 2.0^96,97^.

In secondary analyses of analysis set (i) we also evaluated LDpred^33^. We trained LDpred using HapMap 3 SNPs and using two different LD reference panels: 1000 Genomes project^59^ and UK10K^60^. We used summary statistics from the infinitesimal version of BOLT-LMM (as in SBayesR) and with default parameters, using the parameter --ldr 400. We used the value of “--F” (corresponding to the assumed proportion of causal SNPs, using all the default evaluated values) that yielded the best prediction accuracy in the target sample, yielding anti-conservative accuracy estimates as in P+T.

In analysis sets (ii) and (iii), we trained BOLT-LMM-BBJ, SBayesR-BBJ, and PRS-CS-BBJ (BOLT-LMM, SBayesR, and PRS-CS, respectively, trained using Biobank Japan training data) (average *N*=124K). We selected individuals for training these methods as described in our previous work^13^, but excluding a random subset of 5,000 individuals that were used for evaluating prediction accuracy. For SBayesR-BBJ, we used a subset of individuals (N=50K) from Biobank Japan to compute an in-sample LD, following the recommendations of the authors of SBayesR^38^. For PRS-CS-BBJ, we used the East Asian LD reference panels made publicly available by the authors of PRS-CS (see URLs).

### Loss of accuracy under an infinite European training sample

Under an infinite European training sample, the ratio between 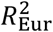 and 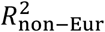, which denote *R*^2^ in an European sample and in a non-European sample, respectively, is approximately given by:

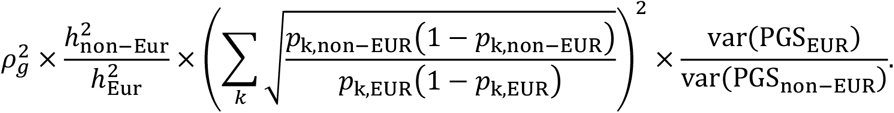

Here, *ρ*_*g*_ is the cross-population genetic correlation, 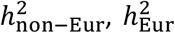 are the heritabilities in the non-European and the European populations, respectively, *k* iterates over causal SNPs, *p*_k,non−EUR_, *p*_k,EUR_ are minor allele frequencies in the non-European and the European population, respectively, and var(PGS_EUR_), var(PGS_non−EUR_) are the variances of the polygenic risk scores in the non-European and the European populations, respectively. This equation is directly derived from Equation 1 in ref.^14^, after assuming that causal SNPs are approximately not in LD with each other, and that the predictor SNPs are the causal SNPs under an infinite sample size.

## Data Availability

PolyPred and PolyPred+ are provided as part of the open-source software package PolyFun, which is freely available at https://github.com/omerwe/polyfun. Access to the UK Biobank resource is available via application (http://www.ukbiobank.ac.uk/). PRS coefficients generated in this study are available for public download at http://data.broadinstitute.org/alkesgroup/polypred_results.

## Acknowledgements

We thank Armin Schoech and Carla Márquez-Luna for helpful discussions. This research was conducted using the UK Biobank Resource under Application #16549 and was funded by NIH grants U01 HG009379, R37 MH107649, R01 MH101244 and R01 HG006399. MK is supported by a Nakajima Foundation Fellowship and the Masason Foundation. WJP is supported by an NWO Veni grant (91619152). ARM is supported by NIMH K99/R00MH117229. HKF is supported by Eric and Wendy Schmidt. AVK is supported by grants 1K08HG010155 and 1U01HG011719 from the National Human Genome Research Institute and a sponsored research agreement from IBM Research. Computational analyses were performed on the O2 High-Performance Compute Cluster at Harvard Medical School.

## URLs

Software implementing PolyPred and PolyPred+: https://www.hsph.harvard.edu/alkes-price/software

Baseline-LF v2.2.UKB annotations and LD-scores for UK Biobank SNPs: https://data.broadinstitute.org/alkesgroup/LDSCORE/baselineLF_v2.2.UKB.tar.gz

Summary LD information of *N*=337K British-ancestry UK Biobank individuals for 2,763 overlapping 3Mb loci: https://data.broadinstitute.org/alkesgroup/UKBB_LD/

PRS coefficients for all analyzed SNPs: https://data.broadinstitute.org/alkesgroup/polypred_results

BOLT-LMM: https://data.broadinstitute.org/alkesgroup/BOLT-LMM

SBayesR: https://cnsgenomics.com/software/gctb

PRS-CS: https://github.com/getian107/PRScs

UK Biobank Resource: http://www.ukbiobank.ac.uk

## Code and data availability

PolyPred and PolyPred+ are provided as part of the open-source software package PolyFun, which is freely available at https://github.com/omerwe/polyfun. Access to the UK Biobank resource is available via application (http://www.ukbiobank.ac.uk). PRS coefficients generated in this study are available for public download at http://data.broadinstitute.org/alkesgroup/polypred_results.

## References

1. Chatterjee, N., Shi, J. & García-Closas, M. Developing and evaluating polygenic risk prediction models for stratified disease prevention. Nat. Rev. Genet. 17, 392–406 (2016).

2. Khera, A. V. et al. Genome-wide polygenic scores for common diseases identify individuals with risk equivalent to monogenic mutations. Nat. Genet. 50, 1219–1224 (2018).

3. Torkamani, A., Wineinger, N. E. & Topol, E. J. The personal and clinical utility of polygenic risk scores. Nat. Rev. Genet. 19, 581–590 (2018).

4. Khera, A. V. et al. Polygenic prediction of weight and obesity trajectories from birth to adulthood. Cell 177, 587–596 (2019).

5. Mavaddat, N. et al. Polygenic risk scores for prediction of breast cancer and breast cancer subtypes. Am. J. Hum. Genet. 104, 21–34 (2019).

6. Li, R., Chen, Y., Ritchie, M. D. & Moore, J. H. Electronic health records and polygenic risk scores for predicting disease risk. Nat. Rev. Genet. 1–10 (2020).

7. Márquez-Luna, C., Loh, P.-R., Consortium, S. A. T. 2 D. (SAT2D), Consortium, S. T. 2 D. & Price, A. L. Multiethnic polygenic risk scores improve risk prediction in diverse populations. Genet. Epidemiol. 41, 811–823 (2017).

8. Grinde, K. E. et al. Generalizing polygenic risk scores from Europeans to Hispanics/Latinos. Genet. Epidemiol. 43, 50–62 (2019).

9. Peterson, R. E. et al. Genome-wide association studies in ancestrally diverse populations: opportunities, methods, pitfalls, and recommendations. Cell 179, 589–603 (2019).

10. Sirugo, G., Williams, S. M. & Tishkoff, S. A. The missing diversity in human genetic studies. Cell 177, 26–31 (2019).

11. Duncan, L. et al. Analysis of polygenic risk score usage and performance in diverse human populations. Nat. Commun. 10, 1–9 (2019).

12. Gurdasani, D., Barroso, I., Zeggini, E. & Sandhu, M. S. Genomics of disease risk in globally diverse populations. Nat. Rev. Genet. 20, 520–535 (2019).

13. Martin, A. R. et al. Clinical use of current polygenic risk scores may exacerbate health disparities. Nat. Genet. 51, 584 (2019).

14. Wang, Y. et al. Theoretical and empirical quantification of the accuracy of polygenic scores in ancestry divergent populations. Nat. Commun. 11, 3865 (2020).

15. Amariuta, T. et al. Improving the trans-ancestry portability of polygenic risk scores by prioritizing variants in predicted cell-type-specific regulatory elements. Nat. Genet. 52, 1346–1354 (2020).

16. Marnetto, D. et al. Ancestry deconvolution and partial polygenic score can improve susceptibility predictions in recently admixed individuals. Nat. Commun. 11, 1–9 (2020).

17. Bitarello, B. D. & Mathieson, I. Polygenic Scores for Height in Admixed Populations. G3 g3.401658.2020 (2020).

18. Chen, M.-H. et al. Trans-ethnic and ancestry-specific blood-cell genetics in 746,667 individuals from 5 global populations. Cell 182, 1198–1213 (2020).

19. Mahajan, A. et al. Trans-ancestry genetic study of type 2 diabetes highlights the power of diverse populations for discovery and translation. medRxiv (2020).

20. Cavazos, T. B. & Witte, J. S. Inclusion of variants discovered from diverse populations improves polygenic risk score transferability. Hum. Genet. Genomics Adv. 2, 100017 (2021).

21. Mills, M. C. & Rahal, C. The GWAS Diversity Monitor tracks diversity by disease in real time. Nat. Genet. 52, 242–243 (2020).

22. Lehmann, B. C., Mackintosh, M., McVean, G. & Holmes, C. C. High trait variability in optimal polygenic prediction strategy within multiple-ancestry cohorts. bioRxiv (2021).

23. Ji, Y. et al. Incorporating European GWAS findings improve polygenic risk prediction accuracy of breast cancer among East Asians. Genet. Epidemiol. (2021).

24. Ruan, Y. et al. Improving Polygenic Prediction in Ancestrally Diverse Populations. medRxiv 2020–12.

25. Cai, M. et al. A unified framework for cross-population trait prediction by leveraging the genetic correlation of polygenic traits. Am. J. Hum. Genet. (2021).

26. Huang, Q. Q. et al. Transferability of genetic loci and polygenic scores for cardiometabolic traits in British Pakistanis and Bangladeshis. medRxiv (2021).

27. Durvasula, A. & Lohmueller, K. E. Negative selection on complex traits limits phenotype prediction accuracy between populations. Am. J. Hum. Genet. 108, 620–631 (2021).

28. Coram, M. A., Fang, H., Candille, S. I., Assimes, T. L. & Tang, H. Leveraging Multi-ethnic Evidence for Risk Assessment of Quantitative Traits in Minority Populations. Am. J. Hum. Genet. 101, 218–226 (2017).

29. Wojcik, G. L. et al. Genetic analyses of diverse populations improves discovery for complex traits. Nature 570, 514–518 (2019).

30. Shi, H. et al. Population-specific causal disease effect sizes in functionally important regions impacted by selection. Nat. Commun. 12, 1–15 (2021).

31. Kuchenbaecker, K. et al. The transferability of lipid loci across African, Asian and European cohorts. Nat. Commun. 10, (2019).

32. Mostafavi, H. et al. Variable prediction accuracy of polygenic scores within an ancestry group. Elife 9, e48376 (2020).

33. Vilhjálmsson, B. J. et al. Modeling Linkage Disequilibrium Increases Accuracy of Polygenic Risk Scores. Am. J. Hum. Genet. 97, 576–592 (2015).

34. Schaid, D. J., Chen, W. & Larson, N. B. From genome-wide associations to candidate causal variants by statistical fine-mapping. Nat Rev Genet 19, 491–504 (2018).

35. Weissbrod, O. et al. Functionally informed fine-mapping and polygenic localization of complex trait heritability. Nat. Genet. 1–9 (2020).

36. Loh, P.-R. et al. Efficient Bayesian mixed-model analysis increases association power in large cohorts. Nat. Genet. 47, 284–290 (2015).

37. Loh, P.-R., Kichaev, G., Gazal, S., Schoech, A. P. & Price, A. L. Mixed-model association for biobank-scale datasets. Nat. Genet. 50, 906–908 (2018).

38. Lloyd-Jones, L. R. et al. Improved polygenic prediction by Bayesian multiple regression on summary statistics. Nat. Commun. 10, 1–11 (2019).

39. Ge, T., Chen, C.-Y., Ni, Y., Feng, Y.-C.A. & Smoller, J. W. Polygenic prediction via Bayesian regression and continuous shrinkage priors. Nat. Commun. 10, 1–10 (2019).

40. Bycroft, C. et al. The UK Biobank resource with deep phenotyping and genomic data. Nature 562, 203 (2018).

41. Nagai, A. et al. Overview of the BioBank Japan Project: study design and profile. J. Epidemiol. 27, S2–S8 (2017).

42. Asiki, G. et al. The general population cohort in rural south-western Uganda: a platform for communicable and non-communicable disease studies. Int. J. Epidemiol. 42, 129–141 (2013).

43. Heckerman, D. et al. Linear mixed model for heritability estimation that explicitly addresses environmental variation. Proc. Natl. Acad. Sci. 113, 7377–7382 (2016).

44. Zeng, J. et al. Signatures of negative selection in the genetic architecture of human complex traits. Nat. Genet. 50, 746 (2018).

45. Gazal, S. et al. Functional architecture of low-frequency variants highlights strength of negative selection across coding and non-coding annotations. Nat. Genet. 50, 1600–1607 (2018).

46. Schoech, A. P. et al. Quantification of frequency-dependent genetic architectures in 25 UK Biobank traits reveals action of negative selection. Nat. Commun. 10, 790 (2019).

47. Duan, S., Zhang, W., Cox, N. J. & Dolan, M. E. FstSNP-HapMap3: a database of SNPs with high population differentiation for HapMap3. Bioinformation 3, 139 (2008).

48. Purcell, S. M. et al. Common polygenic variation contributes to risk of schizophrenia and bipolar disorder. Nature 460, 748–752 (2009).

49. Stahl, E. A. et al. Bayesian inference analyses of the polygenic architecture of rheumatoid arthritis. Nat. Genet. 44, 483–489 (2012).

50. O’Connor, L. J. et al. Extreme Polygenicity of Complex Traits Is Explained by Negative Selection. Am. J. Hum. Genet. 105, 456–476 (2019).

51. Lloyd-Jones, L. GCTB SBayesR shrunk sparse linkage disequilibrium matrices for HM3 variants, summary statistics and predictors generated from ‘Improved polygenic prediction by Bayesian multiple regression on summary statistics’ by Lloyd-Jones, Zeng et al. 2019. (2019) doi:10.5281/ZENODO.3350914.

52. Lam, M. et al. Comparative genetic architectures of schizophrenia in East Asian and European populations. Nat. Genet. 51, 1670–1678 (2019).

53. Nievergelt, C. M. et al. International meta-analysis of PTSD genome-wide association studies identifies sex- and ancestry-specific genetic risk loci. Nat. Commun. 10, 4558 (2019).

54. Sakaue, S. et al. Trans-biobank analysis with 676,000 individuals elucidates the association of polygenic risk scores of complex traits with human lifespan. Nat. Med. 26, 542–548 (2020).

55. Vuckovic, D. et al. The Polygenic and Monogenic Basis of Blood Traits and Diseases. Cell 182, 1214-1231.e11 (2020).

56. Guo, J. et al. Global genetic differentiation of complex traits shaped by natural selection in humans. Nat. Commun. 9, 1–9 (2018).

57. Moser, G. et al. Simultaneous discovery, estimation and prediction analysis of complex traits using a Bayesian mixture model. PLoS Genet 11, e1004969 (2015).

58. Sved, J. A., McRae, A. F. & Visscher, P. M. Divergence between human populations estimated from linkage disequilibrium. Am. J. Hum. Genet. 83, 737–743 (2008).

59. 1000 Genomes Project Consortium. A global reference for human genetic variation. Nature 526, 68–74 (2015).

60. The UK10K Consortium et al. The UK10K project identifies rare variants in health and disease. Nature 526, 82 (2015).

61. Zhang, Q., Privé, F., Vilhjálmsson, B. & Speed, D. Improved genetic prediction of complex traits from individual-level data or summary statistics. Nat. Commun. 12, 4192 (2021).

62. Hu, Y. et al. Leveraging functional annotations in genetic risk prediction for human complex diseases. PLoS Comput. Biol. 13, e1005589 (2017).

63. Marquez-Luna, C. et al. LDpred-funct: incorporating functional priors improves polygenic prediction accuracy in UK Biobank and 23andMe data sets. bioRxiv 375337 (2020).

64. Yang, J., Lee, S. H., Goddard, M. E. & Visscher, P. M. GCTA: a tool for genome-wide complex trait analysis. Am. J. Hum. Genet. 88, 76–82 (2011).

65. Yang, J. et al. Common SNPs explain a large proportion of the heritability for human height. Nat. Genet. 42, 565–569 (2010).

66. HapMap3 Consortium. Integrating common and rare genetic variation in diverse human populations. Nature 467, 52 (2010).

67. Bhangale, T. R., Rieder, M. J. & Nickerson, D. A. Estimating coverage and power for genetic association studies using near-complete variation data. Nat. Genet. 40, 841–843 (2008).

68. Budin-Ljøsne, I. et al. Data sharing in large research consortia: experiences and recommendations from ENGAGE. Eur. J. Hum. Genet. 22, 317–321 (2014).

69. Surakka, I. et al. The impact of low-frequency and rare variants on lipid levels. Nat. Genet. 47, 589–597 (2015).

70. Horikoshi, M. et al. Discovery and fine-mapping of glycaemic and obesity-related trait loci using high-density imputation. PLoS Genet. 11, e1005230 (2015).

71. Pain, O. et al. Evaluation of polygenic prediction methodology within a reference-standardized framework. PLoS Genet. 17, e1009021 (2021).

72. Chung, W. et al. Efficient cross-trait penalized regression increases prediction accuracy in large cohorts using secondary phenotypes. Nat. Commun. 10, 1–11 (2019).

73. Chun, S. et al. Non-parametric Polygenic Risk Prediction via Partitioned GWAS Summary Statistics. Am. J. Hum. Genet. (2020).

74. Im, C. et al. Generalizability of “GWAS Hits” in Clinical Populations: Lessons from Childhood Cancer Survivors. Am. J. Hum. Genet. 107, 636–653 (2020).

75. Daetwyler, H. D., Villanueva, B. & Woolliams, J. A. Accuracy of predicting the genetic risk of disease using a genome-wide approach. PloS One 3, e3395 (2008).

76. Visscher, P. M. & Hill, W. G. The limits of individual identification from sample allele frequencies: theory and statistical analysis. PLoS Genet 5, e1000628 (2009).

77. Wang, G., Sarkar, A., Carbonetto, P. & Stephens, M. A simple new approach to variable selection in regression, with application to genetic fine mapping. J. R. Stat. Soc. Ser. B Stat. Methodol. n/a, (2020).

78. Galinsky, K. J. et al. Estimating cross-population genetic correlations of causal effect sizes. Genet. Epidemiol. 43, 180–188 (2019).

79. Brown, B. C., Ye, C. J., Price, A. L. & Zaitlen, N. Transethnic Genetic-Correlation Estimates from Summary Statistics. Am. J. Hum. Genet. 99, 76–88 (2016).

80. Robinson, M. R. et al. Genotype–covariate interaction effects and the heritability of adult body mass index. Nat. Genet. 49, 1174–1181 (2017).

81. Wray, N. R. et al. Pitfalls of predicting complex traits from SNPs. Nat. Rev. Genet. 14, 507–515 (2013).

82. Kong, A. et al. The nature of nurture: Effects of parental genotypes. Science 359, 424–428 (2018).

83. Bulik-Sullivan, B. K. et al. LD Score regression distinguishes confounding from polygenicity in genome-wide association studies. Nat. Genet. 47, 291–295 (2015).

84. Mak, T. S. H., Porsch, R. M., Choi, S. W., Zhou, X. & Sham, P. C. Polygenic scores via penalized regression on summary statistics. Genet. Epidemiol. 41, 469–480 (2017).

85. Yang, S. & Zhou, X. Accurate and Scalable Construction of Polygenic Scores in Large Biobank Data Sets. Am. J. Hum. Genet. (2020).

86. Qian, J. et al. A fast and scalable framework for large-scale and ultrahigh-dimensional sparse regression with application to the UK Biobank. PLoS Genet. 16, e1009141 (2020).

87. Finucane, H. K. et al. Partitioning heritability by functional annotation using genome-wide association summary statistics. Nat. Genet. 47, 1228–1235 (2015).

88. Purcell, S. et al. PLINK: a tool set for whole-genome association and population-based linkage analyses. Am J Hum Genet 81, 559–75 (2007).

89. Akiyama, M. et al. Characterizing rare and low-frequency height-associated variants in the Japanese population. Nat. Commun. 10, 1–11 (2019).

90. Loh, P.-R. et al. Reference-based phasing using the Haplotype Reference Consortium panel. Nat. Genet. 48, 1443–1448 (2016).

91. Das, S. et al. Next-generation genotype imputation service and methods. Nat. Genet. 48, 1284–1287 (2016).

92. Price, A. L. et al. Principal components analysis corrects for stratification in genome-wide association studies. Nat. Genet. 38, 904–909 (2006).

93. Sakaue, S. et al. A global atlas of genetic associations of 220 deep phenotypes. medRxiv (2020).

94. Gurdasani, D. et al. Uganda genome resource enables insights into population history and genomic discovery in Africa. Cell 179, 984–1002 (2019).

95. Lam, M. et al. RICOPILI: Rapid Imputation for COnsortias PIpeLIne. Bioinformatics 36, 930–933 (2020).

96. Chang, C. C. et al. Second-generation PLINK: rising to the challenge of larger and richer datasets. GigaScience 4, 7 (2015).

97. Purcell, S & Chang, C. PLINK v2.00a3LM.

98. Gazal, S., Marquez-Luna, C., Finucane, H. K. & Price, A. L. Reconciling S-LDSC and LDAK functional enrichment estimates. Nat. Genet. 51, 1202–1204 (2019).

