## Supplementary table captions and Supplementary Note for "Leveraging fine-mapping and non-European training data to improve cross-population polygenic risk scores"

**Supplementary Table 1: Detailed simulation results.** For each combination of generative model parameters, ancestry, and trait we report **(Polygenicity):** trait polygenicity **(European h2):** Trait heritability in Europeans **(Cross-population rg):** Cross-population genetic correlation. **(Method):** The method name (see below). **(#Experiments):** Number of experiments performed. **(Ancestry):** Target ancestry. **(N):** Target ancestry sample size. **(Avg. h2):** Average h2 in the target ancestry. **(s.d. h2):** s.d. h2 in the target ancestry. **(Average R2):** Average $R^2$**.** **(R2 s.e.):** The standard errors of $R^2$. **(R2 (normalized vs. EUR)):** $R^2$ (normalized against the results of the same method in Europeans). **(R2 (normalized vs. EUR) s.e.):** Standard error of $R^2$ (normalized against the results of the same method in Europeans). **(R2 (normalized vs. BOLT-LMM-EUR)):** $R^2$ (normalized vs. BOLT-LMM in non-British Europeans). **(R2 (normalized vs. BOLT-LMM-EUR) s.e.):** The standard error of $R^2$ (normalized vs. BOLT-LMM in non-British Europeans). **(R2 (normalized vs. BOLT-LMM-EUR) P-value):** The p-value of the statistical test testing the null hypothesis that $R^2$ is the same as obtained by BOLT-LMM in non-British Europeans. **(R2 (normalized vs. BOLT-LMM)):** $R^2$ (normalized vs. BOLT-LMM in the target ancestry). **(R2 (normalized vs. BOLT-LMM) s.e.):** The standard error of $R^2$ (normalized vs. BOLT-LMM in the target ancestry). **(R2 (normalized vs. BOLT-LMM) P-value):** The p-value of the statistical test testing the null hypothesis that $R^2$ is the same as obtained by BOLT-LMM in the target ancestry. **(R2 diff vs. BOLT-LMM-EUR):** The difference between the $R^2$ obtained by the current method and BOLT-LMM in non-British Europeans. **(R2 diff vs. BOLT-LMM-EUR s.e.):** The standard error of the difference between the $R^2$ obtained by the current method and BOLT-LMM in non-British Europeans. **(R2 diff vs. BOLT-LMM-EUR P-value):** The p-value of the statistical test testing the null hypothesis that $R^2$ is the same as obtained by BOLT-LMM in non-British Europeans. **(R2 diff vs. BOLT-LMM):** The difference between the $R^2$ obtained by the current method and BOLT-LMM in the target ancestry. **(R2 diff vs. BOLT-LMM s.e.):** The standard error of the difference between the $R^2$ obtained by the current method and BOLT-LMM in the target ancestry. **(R2 diff vs. BOLT-LMM P-value):** The p-value of the statistical test testing the null hypothesis that $R^2$ is the same as obtained by BOLT-LMM in the target ancestry. The method names are as described in the main text, with the following rules. A suffix "-L1" indicates that PolyFun-pred was invoked assuming only one causal SNP per locus (and did not make use of LD information). A suffix "-N" indicates that the training sample size was reduced to the specified number. A suffix "-Nmix" indicates that the target-ancestry training sample size used for estimating mixing weights was modified from 500 to some other number. A suffix "-1000G" indicates that the LD reference panel used European data from the 1000 Genomes project. A suffix "-UK10K" indicates that the LD reference panel used UK10K. A suffix "-HM3" indicates that the SNP set consisted of only 1.2 million HapMap3 SNPs. A suffix "NoFun" indicates that the analysis did not use functional annotations to specify per-SNP prior causal probabilities.

**Supplementary Table 2: Detailed simulation runtime analysis.** For each combination of method and generative model parameters we report (**Method**): method name. (**#Experiments**): number of experiments performed. (**Polygenicity**): trait polygenicity. (**European h2**): h2 in Europeans. (**Average run**

**time (sec)**): average run time in seconds. (**SE (sec)**): the standard error of the runtime in seconds. (**Average run time (hr)**): average run time in hours. (**SE (hr)**): the standard error of the runtime in hours.

**Supplementary Table 3: List of 49 diseases and complex traits.** For each trait, we report its UK Biobank British sample size (used for training most methods), its UK Biobank British heritability estimate and its standard error (estimated using S-LDSC with the Baseline-LF v2.2.UKB model[1]), whether the trait was one of the 7 traits included in the meta-analysis, and whether the trait exists in Biobank Japan.

**Supplementary Table 4: Detailed results of analyses using UKB British training individuals applied to other UKB populations, compared vs. BOLT-LMM.** For each combination of method, ancestry and trait (including meta-analyzed traits) we report (**Method**): The method name (see below). (**N**): Test set sample size; (**R2**): The squared Pearson correlation coefficient between the PRS and the trait after adjusting for covariates (see Methods); (**R2 s.e.**): The standard error of $R^2$, computed via genomic block-jackknife; (**R2 (normalized vs. BOLT-LMM-EUR)**): The $R^2$ value, divided by the $R^2$ of BOLT-LMM in non-British Europeans; (**R2 (normalized vs. BOLT-LMM-EUR) s.e.**): The standard error of the normalized $R^2$; (**R2 diff vs. BOLT-LMM**): The difference between $R^2$ and the $R^2$ obtained by BOLT-LMM; (**R2 diff vs. BOLT-LMM s.e.**) The standard error of the difference between $R^2$ and the $R^2$ obtained by BOLT-LMM, computed via genomic block-jackknife; (**R2 diff vs. BOLT-LMM (normalized vs. BOLT-LMM-EUR)**): The difference between normalized $R^2$ and the normalized $R^2$ obtained by BOLT-LMM; (**R2 diff vs. BOLT-LMM (normalized vs. BOLT-LMM-EUR) s.e.**): The standard error of the difference between normalized $R^2$ and the normalized $R^2$ obtained by BOLT-LMM; (**R2 vs. BOLT-LMM (normalized vs. BOLT-LMM-EUR) P-value**): The P-value of the difference between the normalized $R^2$ and the normalized $R^2$ obtained by BOLT-LMM; (**Regression slope**): Slope obtained when regressing the true phenotype on the PRS; (**Regression slope s.e.**): The standard error of the regression slope, computed via genomic block-jackknife; (**R2 ind-s.e.**): The standard error of $R^2$, computed via jackknife over individuals; (**Mixing weights**): The mixing weights of combined methods (blank for non-combined methods). The first value is the intercept, and the other values are PolyPred, BOLT-LMM (resp. SBayesR or PRS-CS), and BOLT-LMM-BBJ (when there are four numbers) (resp. SBayesR-BBJ or PRS-CS-BBJ); (**P vs. Europeans**): The p-value of the hypothesis that the R2 obtained in the target ancestry is the same as in non-British Europeans (under the same method and trait), as computed via a conservative Wald test (Methods). The method names that are not explicitly defined in the main text are the following: **Methods ending with -pX** (where X is a number) are methods using a fixed mixing weight X for PolyPred and its extensions; **Methods ending with with -100** use 100 individuals from the target cohort to estimate mixing weights (instead of 500 as used by most combined methods); **BOLT-LMM-727K**: BOLT-LMM using only genotyped SNPs; **LDpred-1000G-p**: LDpred using the 1000 genomes project Europeans as an LD reference panel, and assuming that proportion p of causal SNPs are causal; **LDpred-1000G-cheat**: LDpred using the 1000 genomes Europeans as an LD reference panel, and using the best value of p for each trait (as determined via $R^2$ in the test set); **LDpred-UK10K-p**: LDpred using the UK10K cohort as an LD reference panel, and assuming that proportion p of causal SNPs are causal; **LDpred-UK10K-cheat**: LDpred using the UK10 cohort as an LD reference panel, and using the best value of p for each trait (as determined via $R^2$ in the test set) (we caution that standard errors of methods using only PIP>0.95 SNPs may not be accurate because of the small number of SNPs used); **PRS-CS-phi0.0001**: PRS-CS with –phi=0.0001; **PRS-CS-phi0.01**: PRS-CS with –phi=0.01; **PRS-CS**: PRS-CS using a Biobank reference panel LD, and without specifying –phi; **PRS-CS-cheat**: PRS-CS that uses the best value of –phi for each target ancestry; **PRS-CS-1000G**: PRS-CS, using $N$=489 1000 Genomes project Europeans as an LD reference panel; **PolyFun-pred-pipP**: PolyFun-pred restricted to SNPs with PIP greater than P; **PolyFun-pred-NoFun**:

PolyFun-pred without using functional annotations; **PRS-CS-BBJ**: PRS-CS, trained on Biobank Japan individuals (using a UK Biobank East-Asian LD reference panel); **P+T-pX**: P+T that uses only SNPs with BOLT-LMM P-value <X; **P+T-cheat**: P+T that uses the best value of X for each target ancestry. **PolyPred+-Ext**: PolyPred+ with mixing weights estimated in Biobank Japan; **PolyPred-pipP**: PolyPred restricted to SNPs with PIP greater than P; **PolyPred-NoFun**: PolyPred without using functional annotations; **SBayesR-2.8M**: SBayesR using 2.8M common SNPs selected by the SBayesR authors; **SBayesR-UK10K:** SBayesR, using UK10K (*N*=3000) as an LD reference panel; **SBayesR-1000G**: SBayesR, using the 1000 Genomes project Europeans (N=489) as an LD reference panel; SBayesR-UK10K-489: SBayesR, using a subset of UK10K individuals matched to the 1000 Genomes project Europeans sample size (N=489) as an LD reference panel; **PolyFun-pred-UK10K**: PolyFun-pred, using UK10K (*N*=3000) as an LD reference panel; **BOLT-LMM-N-African**: BOLT-LMM, evaluated by subsampling the test set of each population to the African sample size of the corresponding trait (unless the sample size was smaller than the African sample size). To see the numerical results of the analyses reported in the main text, please filter the **Trait** column to show only the trait 'Meta-Analysis'.

**Supplementary Table 5: Comparisons between pairs of methods in analyses of real UK Biobank and Biobank Japan traits.** The table reports comparisons of selected pairs of methods mentioned in the main text. For each pair of methods we report its training data (UKB indicates individual-level data or summary statistics from 337K UK Biobank British individuals; ENGAGE indicates summary statistics from the European Network for Genetic and Genomic Epidemiology; UKB+BBJ indicates a combination of UK Biobank and Biobank Japan training data); the names of the two methods (Method1 and Method2); the target ancestry; the trait name; the target ancestry sample size; the accuracy ($R^2$) of method1; the difference in $R^2$ between the two methods (Method1 $R^2$ – Method2 $R^2$) and its standard error; and the p-value of the difference, as computed via a genomic block jackknife over 200 genomic blocks.

**Supplementary Table 6: Detailed results of analyses using UKB British training individuals applied to other UKB populations, compared vs. PolyPred.** The table is analogous to Table 4, but all results are normalized and compared with respect to PolyPred instead of BOLT-LMM.

**Supplementary Table 7: Ancestry-specific SNP heritability estimates in the UK Biobank, across 7 independent complex traits.** For each trait (including meta-analyzed traits) we report its sample size (n), its SNP heritability estimate (h2g) and its standard error (se). All estimates were performed using GCTA[2] with HapMap 3[3] SNPs due to the relatively small sample sizes. Non-British Europeans were down-sampled to 10,000 individuals to facilitate the analysis. Meta-analyzed h2g was computed via the average h2g, and the meta-analyzed standard error was computed via the square root of the average sampling variance, divided by the square root of the number of traits.

**Supplementary Table 8: Prediction accuracy using summary statistics from the from the European Network for Genetic and Genomic Epidemiology.** The table is analogous to Supplementary Table 4, but reports results based on training data from the European Network for Genetic and Genomic Epidemiology (ENGAGE) for four traits (BMI, waist-hip-ratio (adjusted for BMI), total cholesterol, and triglycerides) (average *N*=61K) instead of training data based on up to *N*=337K UK Biobank British individuals.

**Supplementary Table 9: Detailed results of analyses applied to Biobank Japan and to Uganda-APCDR.** The table is analogous to Table 4, but includes columns comparing each method to PolyPred in addition to columns comparing each method to BOLT-LMM.

**Supplementary Table 10: Comparing prediction accuracy in UK Biobank Non-British Europeans and in Biobank Japan when using equal training set sample sizes.** For each of 7 independent traits we report (**N**) its Biobank Japan training sample size (which was also used for the UK Biobank British training sample size in this analysis); (**h2g (UKB-EUR)**) its non-British European SNP heritability, as estimated by BOLT-REML; (**h2g (BBJ)**) its Biobank Japan SNP heritability, as estimated by BOLT-REML; (**R2-expected (UKB EUR)**) the expected $R^2$ in non-British Europeans as a function of training set sample size and SNP heritability, based on theory (see Supplementary Note); (**R2-expected (Biobank Japan)**) the expected $R^2$ in Biobank Japan as a function of training set sample size and SNP heritability, based on theory; (**R2 (UKB-EUR)**) the $R^2$ obtained in practice in non-British Europeans when training BOLT-LMM using a UK Biobank British training sample with the same sample size as the Biobank Japan training sample size; (**R2 (BBJ)**) the $R^2$ obtained in practice in 5K Biobank Japan individuals when training BOLT-LMM using a Biobank Japan training sample.

**Supplementary Table 11: Description of 187 baseline-LF model annotations used by PolyFun-pred**. For each annotation we report #SNPs in the annotation (unless it is a continuous-valued annotation), #common (MAF>0.05) SNPs in the annotation, whether it is binary or continuous-valued, and a literature reference.

### Supplementary Note

#### Simulations with reference LD

The simulations described in the main text use in-sample LD (i.e., LD summary data based on the UK Biobank GWAS sample). However, researchers often do not have access to in-sample LD, necessitating external LD reference panels. We thus evaluated modified versions of PolyFun-pred, SBayesR and PRS-CS that use summary LD estimated from 1000 Genomes project Europeans[4] ($N$=489). We note that this LD reference panel is both smaller than the UK Biobank British LD reference panel ($N$=337K) and less well-matched to the GWAS sample, because it consists of pan-European ancestries rather than only British-ancestry individuals. We excluded BOLT-LMM from these analyses because it requires individual-level data.

The results of simulations with reference LD are reported in Supplementary Table 1. All methods became less accurate when using 1000 Genomes project Europeans LD summary data. The loss of accuracy was modest for SBayesR (-5% $R^2$ for non-British Europeans vs. using in-sample LD) but severe for PRS-CS (-42% $R^2$ for non-British Europeans vs. using in-sample LD) and PolyFun-pred (-90% for non-British Europeans vs. using in-sample LD). We caution that the differences observed in real trait analysis for SBayesR and PRS-CS were substantially different from those observed in our simulations (large loss of accuracy for SBayesR, no significant loss of accuracy for PRS-CS), suggesting that the effect of LD mismatch on PRS accuracy may be sensitive to the underlying genetic architecture.

We performed 3 secondary analyses. First, we evaluated a modified version of PolyFun-pred that uses summary LD from UK10K[5] ($N$=3,567). We observed only a moderate loss of accuracy in PolyFun-pred vs. using in-sample LD (-8% $R^2$ in non-British Europeans) (Supplementary Table 1). However, we caution that using UK10K led to substantial and statistically significant loss of accuracy in real trait analysis, suggesting that the results may be sensitive to the underlying genetic architecture. Second, we evaluated modified versions of PolyFun-pred using subsets of UK10K as an LD reference panel, ranging from *N*=3,000 to *N*=489 (matching the 1000 Genomes project Europeans reference LD sample size). The accuracy of PolyFun-pred decreased with the LD reference panel sample size, with the loss in accuracy vs. using in-sample LD (for non-British Europeans) ranging from -8% for *N*=3,000, to -90% for *N*=489 (Supplementary Table 1). Finally, we evaluated a modified version of PolyFun-pred (PolyFun-pred1) that assumes a single causal variant per locus, precluding the need for a reference LD panel (because fine-mapping under a single casual variant assumption does not require any LD information[1]). PolyFun-pred1 was substantially less accurate than all other methods (including P+T) and is thus not recommended for polygenic prediction (Supplementary Table 1).

We conclude that the accuracy of all methods increases with the size of the LD reference panel and its concordance with the GWAS sample population, but that the relationship may depend on the underlying genetic architecture. Hence, it may be best to assess the accuracy obtained under various LD reference panels using real trait analysis rather than simulations. Specifically, the simulation results do not support the use of PolyPred-S or PolyPred-P in the specific scenarios considered in these simulations. However, real data results with very large LD reference panels do support the use of PolyPred-S or PolyPred-P (Figure 5). We did not perform simulations with very large unmatched LD (analogous to Figure 5), as this would have required another very large individual-level data set in addition to UK Biobank.

#### Causal vs. tagging effects

We consider a linear model $y = \sum_i x_i \beta_i + \epsilon$ where $y$ is a trait, $x_i$ is the number of minor alleles at SNP $i$, $\beta_i$ is the (true) causal effect size of SNP $i$, and $\epsilon$ is a residual term sampled from a normal distribution. We consider a method (such as PolyFun-pred) that estimates $\beta_i$. If the generative model holds and all SNPs $i$ are considered in the estimation procedure, then the estimated value $\hat{\beta}_i$ is a consistent estimator of $\beta_i$, and thus $\hat{\beta}_i$ represents a causal effect. In contrast, if only a subset of the SNPs, such as HapMap3 SNPs, are considered in the estimation procedure (i.e. if we incorrectly assume the generative model $y = \sum_{i \in S} x_i \beta_i + \epsilon$, where $S$ is a subset of SNPs) then the estimated value $\hat{\beta}_i$ represents a linear combination of $\beta_i$ and of the effect sizes of other SNPs.

The exact value estimated by $\hat{\beta}_i$ depends on the estimation procedure. For example, assuming an ordinary least squares estimator for simplicity, the vector $\widehat{\boldsymbol{\beta}}_S$ of estimated coefficients is a consistent estimator of $\left[I_{m-k}\ R_{SS}^{-\mathbf{1}} R_{S\bar{S}}\right]\boldsymbol{\beta}$, where $m$ is the total number of SNPs, $k$ is the number of SNPs in the set $S$, $R_{SS}$ is the LD matrix of the SNPs in the set $S$, $R_{S\bar{S}}$ is a matrix wherein each entry $i, j$ is the correlation between SNP $i$ in the set $S$ and SNP $j$ in the set of SNPs that are not in $S$, and $\boldsymbol{\beta}$ is the vector of true effect sizes, assuming without loss of generality that the set $S$ includes the first $k$ SNPs (out of $m$ SNPs considered). It is easy to derive this quantity by writing down the conditional expectation of $\widehat{\boldsymbol{\beta}}_S$ under an ordinary least squares estimator, given by $\mathrm{E}\left[\widehat{\boldsymbol{\beta}}_S \mid \boldsymbol{\beta}\right] = \mathrm{E}[(\boldsymbol{X}_S^T \boldsymbol{X}_S)^{-1} \boldsymbol{X}_S^T \boldsymbol{y} \mid \boldsymbol{\beta}]$, where $\boldsymbol{y} = \boldsymbol{X}\boldsymbol{\beta} + \boldsymbol{\epsilon}$ is a vector of observed phenotypes and $\boldsymbol{X}$ is the corresponding matrix of SNPs, $\boldsymbol{X}_S$ is the submatrix of **X** consisting of columns of SNPs in the set $S$, and we assume that $\epsilon$ is independent of $\boldsymbol{X}$.

#### Investigating if off-cohort loss of accuracy is driven by SNP heritability differences

We investigated if lower prediction accuracies in Biobank Japan vs. the UK Biobank can be largely explained by SNP heritability differences. We began by comparing trait heritabilities across the UK Biobank and Biobank Japan, using BOLT-REML[6] applied to UK Biobank British-ancestry individuals (average *N*=325K) and to Biobank Japan (average *N*=124K), restricting to HapMap 3 SNPs. The average heritability in the UK Biobank was 67% larger (Supplementary Table 10), indicating differences in either trait measurement, cohort ascertainment, the ability of HapMap 3 SNPs to tag East Asian causal SNPs[7], or in the true underlying heritabilities (we could not perform a similar analysis with UK Biobank East Asian individuals due to small sample sizes leading to large standard errors). We next asked if the observed differences in PRS accuracy between Biobank Japan and the UK Biobank can be explained by the 67% increased average SNP heritability in the UK Biobank. To this end, we computed the expected $R^2$ within each cohort as function of SNP heritability, sample size, and the effective number of independent SNPs[8,9]:

$$\mathrm{E}[R^2] = h^2 \frac{h^2}{h^2 + \frac{m}{n}}.$$

Here, $h^2$ is SNP heritability, $n$ is sample size, and $m$ is the effective number of independent SNPs (which we specified as 55,000, determined by dividing the number of HapMap 3 SNPs by their average within-HapMap 3 LD-score). We used the smaller Biobank Japan sample size in both cohorts to eliminate differences due to sample size differences (by choosing a random subset of UK Biobank British individuals as a training set). The average expected $R^2$ in the UK Biobank was 104% larger than in Biobank Japan (Supplementary Table 10). We then trained BOLT-LMM using subsets of the UK Biobank British sample (matching the Biobank Japan sample size for each trait) and applied the predictions to UK Biobank non-British Europeans. The average $R^2$ in UK Biobank non-British Europeans (when training BOLT-LMM using the reduced British training sample) was 108% larger than the average $R^2$ in Biobank Japan (when training BOLT-LMM using the Biobank Japan training sample) (Supplementary Table 10), strongly consistent with the 104% increase expected from theory. Finally, we determined that when training BOLT-LMM using the full UK Biobank British training set (average *N*=325K), the average $R^2$ in UK Biobank East Asians across the 7 independent traits is 93% larger than in Biobank Japan (Supplementary Tables 4 and 9), broadly consistent with the previous results. Assuming that the main factor differentiating the UK Biobank East Asian sample from the Biobank Japan sample is SNP heritability differences (rather than differences in MAF, LD, or causal effect sizes), these findings suggest that the main factor leading to lower prediction accuracies in Biobank Japan vs. the UK Biobank is SNP heritability differences.

To further investigate if off-cohort loss of accuracy is driven by SNP heritability differences, we compared prediction accuracies in UK Biobank East Asians and in Biobank Japan, when training BOLT-LMM using the Biobank Japan training sample. The average relative-$R^2$ in UK Biobank East Asians across the 7 independent traits was 9.0% larger (Supplementary Tables 4,10), though the difference was not statistically significant (P=0.18), possibly owing to the small UK Biobank East Asian sample size.

Although these results are not conclusive, they suggest that heritability differences drive most of the differences in prediction accuracies observed between the UK Biobank and Biobank Japan. Surprisingly, these results are consistent with a model in which HapMap 3 SNPs in Biobank Japan tag approximately 50% of the causal SNPs that they tag in the UK Biobank, rather than a model in which SNP heritabilities in Biobank Japan are smaller due to smaller causal effect sizes. This is because under the second model, we would expect to see large increase in prediction accuracy in UK Biobank East Asians vs. Biobank Japan when training BOLT-LMM using Biobank Japan (compared with only a 9.0% increase observed in practice). A partial explanation is that the HapMap 3 SNP set consists of a combination of two genotyping chips, one of which is explicitly designed to optimize tagging in Europeans[10].

#### Decomposing the PolyFun-pred and BOLT-LMM predictors into shared and non-shared components

A linear combination of PRS predictors is not necessarily suboptimal, even if the methods are correlated. (As an extreme example, a linear combination of two perfectly correlated predictors is optimal.) However, a linear combination could be suboptimal if the correlation between the (effect sizes underlying the) two predictors varies across the genome. As an extreme example, consider a scenario where one predictor is perfectly accurate across the first half of a chromosome but uninformative across the second half, whereas the second predictor is uninformative across the first half but perfectly accurate across the second half. Clearly, the optimal combination would use only the (effect sizes of the) first predictor for the first half of the chromosome, and only the (effect sizes of the) second predictor for the second half of the chromosome. However, a simple linear combination assigns only a single mixing weight to each predictor, and will thus assign equal weights to both predictors, resulting in a suboptimal predictor.

We performed several attempts to improve upon a simple linear combination of PRS predictors by partitioning the genome into segments and estimating different linear mixing weights in different segments. However, this more complex approach did not outperform the simple approach of assigning a simple mixing weight to each predictor (results not shown), and we thus did not pursue it further.

### References


1. Weissbrod, O. *et al.* Functionally informed fine-mapping and polygenic localization of complex trait heritability. *Nat. Genet.* 1–9 (2020).
2. Yang, J., Lee, S. H., Goddard, M. E. & Visscher, P. M. GCTA: a tool for genome-wide complex trait analysis. *Am. J. Hum. Genet.* **88**, 76–82 (2011).
3. HapMap3 Consortium. Integrating common and rare genetic variation in diverse human populations. *Nature* **467**, 52 (2010).
4. 1000 Genomes Project Consortium. A global reference for human genetic variation. *Nature* **526**, 68–74 (2015).
5. The UK10K Consortium *et al.* The UK10K project identifies rare variants in health and disease. *Nature* **526**, 82 (2015).
6. Loh, P.-R. *et al.* Contrasting genetic architectures of schizophrenia and other complex diseases using fast variance-components analysis. *Nat. Genet.* **47**, 1385–1392 (2015).

7. Bhangale, T. R., Rieder, M. J. & Nickerson, D. A. Estimating coverage and power for genetic association studies using near-complete variation data. *Nat. Genet.* **40**, 841–843 (2008).

8. Daetwyler, H. D., Villanueva, B. & Woolliams, J. A. Accuracy of predicting the genetic risk of disease using a genome-wide approach. *PloS One* **3**, e3395 (2008).

9. Visscher, P. M. & Hill, W. G. The limits of individual identification from sample allele frequencies: theory and statistical analysis. *PLoS Genet* **5**, e1000628 (2009).

10. Duan, S., Zhang, W., Cox, N. J. & Dolan, M. E. FstSNP-HapMap3: a database of SNPs with high population differentiation for HapMap3. *Bioinformation* **3**, 139 (2008).
